# Improving Health Service Utilization Through Proactive Community Case Management in Côte d’Ivoire: A Controlled Interrupted Time Series Analysis

**DOI:** 10.64898/2026.11.26.26369076

**Authors:** Arsène Brunelle Sandie, Brehima Togola, Samba Mamadou, Lamine Guindo, Allassane Foungoye Ouattara, Youssouf Keita, Richard Yapi, Tom Spears, Jeanne d’Arc Koffi-Kouakou, Amadou Beydi Cisse, Christian Rusangwa, Jessica Beckerman, Emily Treleaven, Simon Cousens, Maud Amon Akissi Tanoh, Ari Johnson

## Abstract

**Introduction:** Use of reproductive, maternal, child and adult healthcare remains low in rural Côte d’Ivoire. Professional community health workers, strengthened primary care and user fee removal each reduce barriers to care and, combined, could increase use. We assessed their combined effect, delivered as Proactive Community Case Management (ProCCM), in Madinani, a rural district.

**Methods:** We conducted a controlled interrupted time series analysis of District Health Information System data (DHIS-2), 2019-2024. Madinani was the intervention district; 20 comparison districts were selected from the 101 eligible districts as those whose deseasonalised pre-intervention trajectory for the primary outcome lay closest to Madinani’s. The primary outcome was monthly curative consultations among children under five. Multilevel negative binomial models with a district random intercept estimated level and trend changes. Eligible districts were ranked by their post-to pre-intervention ratio.

**Results:** Facility-based all-cause under-five consultations rose (ratio of rate ratios (RRR) 2.00, 95% CI 1.55 to 2.57; 1.71, 1.37 to 2.13 under a common pre-intervention slope). With facility and community care combined, treatment increased for diarrhoea with oral rehydration solution and zinc (8.37, 5.29 to 13.24), pneumonia with antibiotics (4.97, 3.34 to 7.41) and malaria with artemisinin-based combination therapy (1.87, 1.32 to 2.66). All-age curative consultations more than doubled (2.20, 1.76 to 2.74) and third-dose intermittent preventive treatment in pregnancy increased (1.36, 1.06 to 1.75). There was no evidence of an effect on institutional deliveries or postnatal care. Madinani’s post:pre ratio for facility-based under-five consultations ranked fifth of 102 districts (p=0.049).

**Conclusions:** Combining user fee removal, professional CHWs, and upgraded primary care was associated with increased use of care by children and people of all ages in Côte d’Ivoire. Policymakers could consider this model to increase access to care.

**What is already known on this topic:**

- Professional Community Health Workers, user-fee removal, and upgraded primary care have each been shown to improve access to care and health outcomes.
- Prior research on combining these interventions, through Proactive Community Case Management, has demonstrated large and rapid improvements in access to care, maternal healthcare outcomes, and child survival in pre-post studies.
- Further research is needed to assess the impact of ProCCM relative to a counterfactual in controlled studies.

**What this study adds:**

- The current study uses a controlled interrupted time series methodology to quantify the impact of ProCCM on access to care.
- By comparing the ProCCM district with a comparison group matched on pre-intervention trajectory, with its own history, and with every other district in the country, the study found that ProCCM increased use of care among children and patients of all ages and quantifies how much of that estimate rests on the assumption that the two arms would otherwise have followed parallel trends.

**How this study might affect research, practice or policy:**

- The findings of this study suggest that Ivorian public health policymakers should consider investment in professional CHWs, user fee removal, and primary care strengthening to increase healthcare service utilization.

## INTRODUCTION

Côte d’Ivoire, like most sub-Saharan African (SSA) countries, faces challenges of low access to reproductive, maternal, neonatal, and child health (RMNCH) services. This is reflected by relatively high rates of under-five mortality (U5M) and maternal deaths, estimated at 74 deaths per 1,000 live births and 385 deaths per 100,000 live births, respectively (1). U5M is higher in rural (84 per 1,000 live births) than in urban areas (64 per 1,000).

There is limited literature on barriers to accessing RMNCH services in rural settings of Côte d’Ivoire. However, in similar settings elsewhere, various structural barriers interact to delay or prevent access to RMNCH services in rural areas. These include geographic and transportation challenges in remote areas (2–4); health systems and health policy constraints, together with gaps in health governance that accentuate challenges in the availability and readiness of health services, understaffing, and lack of skilled providers in rural areas (5, 6). Integrated Community Case Management (ICCM) was developed by WHO and UNICEF to improve prompt access to quality care for children in rural and hard-to-reach areas (7). It consists of training, supporting, and supplying community health workers (CHWs) to provide diagnostic, treatment, and referral services for malaria, pneumonia, and diarrhea (8). ICCM decreases treatment delays and addresses inequity in health service access for children (9). A five-country study found ICCM significantly reduced under-five mortality (10). Côte d’Ivoire and many other SSA countries have adopted ICCM within their national community health strategies (11, 12).

However, impacts documented in trials have not been consistently reproduced at scale: gaps in delivery design leave persistent financial, geographic and quality barriers to care (13–15). Implementation also faces weak supervision and mentorship, inadequate CHW remuneration and motivation, and poor supply chain management (16). Low utilisation in Ethiopia reduced its impact (17). Gaps in CHW knowledge of assessing and treating common childhood illness are also documented (18).

To address these ICCM gaps, Muso (19) proposed a Proactive Community Case Management (ProCCM) health system strengthening intervention (20). ProCCM seeks to mitigate geographical, economic, quality, and acceptability barriers to rapid healthcare access for children and women. In Yirimadio, peri-urban Mali, a seven-year before-and-after study documented improved early access to malaria treatment and a fall in under-five mortality from 154 to 7 deaths per 1,000 live births (20).

A cluster-randomized trial of the ProCCM intervention conducted in Bankass, a rural conflict-affected area in Mali, documented a 63% reduction from baseline in the under-five mortality rate across intervention and control arms during the trial (21), with no difference between arms (proactive CHW home visits versus care from CHWs at a community-based fixed site). The process evaluation attributed the decline in both arms to user fee removal, the availability of professional CHWs in every village, and improved quality of care at primary health clinics (22).

Since December 2022 the MoHPHUHC, with Muso, has implemented ProCCM in Madinani, a health district comprising 25 catchment areas at launch. This study forms part of a larger controlled before-and-after study with a waitlist design and uses controlled interrupted time series analysis of routine data to assess its effectiveness (23). The findings could inform MoHPHUHC investment priorities for maternal health, child survival and universal health coverage.

## METHODS

### Study setting

Madinani health district, in the Denguélé region about 700 km north-west of Abidjan, was selected by Muso with the MoHPHUHC on pre-defined criteria including high under-five mortality, low access to healthcare and absence of competing interventions. It has an estimated 106 000 inhabitants across roughly 7 000 km², and 25 government-owned facilities: two district hospitals, 13 community health centres and 10 rural health posts, the latter two categories forming the primary healthcare level. Some villages lie nearly 80 km from their nearest facility, and no formal private provider reports into DHIS-2, so these data capture all formal care in the district essentially. Further detail on the setting is given in the online supplement.

### Intervention

Community members were recruited, trained over about 36 days, and accredited by examination as proactive professional CHWs (ProCHWs), salaried at 60 000 FCFA per month (about US$107); government CHWs outside ProCCM are unsalaried and receive 60 000 FCFA quarterly. ProCCM has five components. First, proactive case detection through home visits by ProCHWs, targeting two visits per household per month. Second, doorstep care, delivered both at those visits and on demand, since ProCHWs live in the villages they serve: counselling, diagnosis and treatment, with ProCHWs equipped with malaria rapid diagnostic tests, respiratory rate timers and mid-upper-arm-circumference tapes, carrying artemisinin-based combination therapy, oral rehydration salts and zinc and dispersible amoxicillin, and referring or evacuating under iCCM protocols. Third, dedicated supervision, in which trained supervisors observe ProCHWs during service delivery, meet patients without the ProCHW present to collect feedback, and hold monthly individual and weekly group sessions. Fourth, removal of user fees at both community and facility level. Fifth, strengthening of primary healthcare centres through infrastructure improvements, clinical training, mentorship and systems strengthening. In the literature, ProCCM has sometimes denoted proactivity alone rather than professionalization (24–26).

### Study design

We used a controlled interrupted time series design over 47 pre-intervention months (January 2019 to November 2022) and 24 post-intervention months (January 2023 to December 2024); December 2022, the month of launch, was excluded as a transition month. Madinani was the intervention unit. Abidjan districts and Adzopé, which contains a catchment area also piloting ProCCM, were excluded, leaving 101 eligible comparison districts. The initial plan was to use all eligible districts as the comparison group, but their pre-intervention trend differed from Madinani’s (Supplementary Figure S1). We therefore selected the comparison districts whose pre-intervention trajectory was closest to Madinani’s. Twenty districts were selected by a rule applied to the primary outcome alone and without reference to the post-intervention period: each district’s log rate was deseasonalised, centred on its own pre-intervention mean and compared month by month with Madinani’s. The 20 districts with the smallest root mean square distance were retained. Matching on the shape of the trajectory rather than its level means comparison districts need only share Madinani’s pattern of change. The group size was fixed at 20 before any effect was estimated. The single comparison line in the figures is the mean of the 20 districts. Community activity in comparison districts is delivered by the national iCCM programme, whose volunteer CHWs are unsalaried and received no support from Muso.

Parallel trends were tested for every outcome by fitting, to the pre-intervention period alone, a model with calendar-month effects, centred calendar time, study arm and their interaction. The primary model gives each arm its own pre-intervention slope and projects it forward, so its counterfactual assumes that Madinani’s pre-intervention trend relative to the comparison districts would have continued. If that divergence was transient, the projection biases the estimated effect: away from the null where Madinani was declining faster than its comparators, and towards it where Madinani was rising faster. Every outcome was therefore also fitted with a single slope common to both arms, which instead treats any pre-intervention divergence in trend as chance variation. Both estimates are reported; neither is correct a priori, and the distance between them measures how much each estimate depends on that assumption.

### Data source

Data were extracted from the District Health Information System (DHIS-2), the national platform for the Health Monitoring Information System in Côte d’Ivoire, for January 2019 to December 2024.

### Outcomes

#### Primary outcome

The primary outcome was primary healthcare utilisation among children aged 0-59 months, measured as the monthly number of all-cause curative consultations and constructed separately from facility records, from ProCHW records, and from the two combined. The monthly count was the dependent variable, with the logarithm of the under-five population as an offset.

#### Secondary outcomes

Secondary outcomes were utilisation among women aged 15-49 years and in the general population and, among children aged 0-59 months, confirmed malaria treated with ACT, diarrhoea treated with ORS and zinc, and pneumonia treated with antibiotics. Each was analysed with two denominators: per 1,000 children under five, measuring utilisation, and per 1,000 under-five consultations, measuring case mix. The second is itself affected by the intervention, so a fall in the case-mix version is compatible with no change in the number treated; both are reported. Maternal indicators were first antenatal visit in the first trimester and third-dose IPTp-SP (per 1,000 expected pregnancies), institutional deliveries and postnatal care within 72 hours (per 1,000 expected live births), and new contraceptive users (per 1,000 women aged 15-49). For the general population we used curative consultations for all causes and ages per 1,000 total population. Denominators were the projected subnational populations from DHIS-2.

### Data management

Data were managed at facility-month level, before aggregation. Extreme values were identified by a robust within-facility rule on the log scale, requiring a value to be extreme both against the facility’s own median absolute deviation and on an interpretable scale relative to its median; 0.6% to 2.0% of facility-months were flagged, and medians were computed separately by period in every district so that both arms were screened symmetrically (27, 28). Flagged values were set to missing and, with genuinely missing facility-months, imputed by predictive mean matching with five donors within district, using calendar month, calendar time, period, the facility’s own mean of the variable, its caseload, and its reported completeness of returns (29). Twenty imputed datasets were generated and combined by Rubin’s rules with Barnard-Rubin degrees of freedom (30). A second dataset retained only facilities reporting at least 90% of months with no gap longer than three months, used as a completeness sensitivity analysis. The full rule and imputation model are given in the online supplement.

### Analytical approach and modelling

All outcomes were analysed by multilevel mixed-effects negative binomial regression with a district random intercept, fitted to the monthly count with the logarithm of the denominator as an offset, so exponentiated coefficients are rate ratios. The effect of ProCCM is the exponentiated arm-by-period interaction, a ratio of rate ratios (RRR): the change in Madinani’s rate at launch, relative to the rate projected from its pre-intervention trend, divided by the corresponding change in the comparison districts. It is the multiplicative counterpart of a difference-in-differences estimate, described as a ratio in ratios by Lee and Lee (31). Denominators expressed as annual expected counts were divided by 12, a rescaling that leaves every rate ratio unchanged.

The model contains a district random intercept; calendar-month effects; an indicator for study arm; calendar time interacted with arm, allowing different pre-intervention trends; an indicator for the post-intervention period, as a main effect and interacted with arm, the interaction being the step change attributable to ProCCM; and time since launch, likewise, allowing the effect to grow or decay. Entering the post-intervention terms as main effects keeps any national change in January 2023 in the comparison arm. The second specification omits the arm-by-calendar-time interaction. We also report the combined effect 24 months after launch, obtained as a linear combination of the two coefficients so that its confidence interval accounts for their covariance rather than treating them as independent.

Pre- and post-intervention distributions in each arm are shown as box plots (Supplementary Figures S2-S6).

As a second, model-free summary that uses each eligible district individually, we computed for every district the ratio of its mean monthly value over the 24 post-intervention months to its mean monthly value over the 47 pre-intervention months and examined where Madinani ranked within the resulting distribution of ratios, rank 1 being the largest relative increase: 102 districts for facility-based outcomes, 99 for community and combined outcomes. Under the null that Madinani is exchangeable with the others its rank is uniform, so rank divided by the number of districts is a one-sided empirical p-value, free of modelling assumptions. All eligible districts were ranked, rather than only the 20 matched comparators, so that this check does not depend on the matching step or on the choice of group size; against 20 comparators the smallest attainable p-value would be 1/21 (0.048).

Analyses used facility, community and combined facility and community data. All analyses were conducted in Stata version 19.5 (32).

### Sensitivity analysis

#### Five sensitivity analyses were specified in advance: varying the comparison group from 5 to

101 districts; re-selecting on pre-intervention level and slope rather than trajectory; re-selecting separately for each outcome on its own pre-intervention series, with the primary-outcome set retained for the main analysis; repeating the analysis on the nearly complete dataset; and fitting every outcome under both trend specifications. A sixth, added after the others, gave every district its own random slope on calendar time, with Madinani’s slope either estimated separately or partially pooled with the comparison districts’ slopes; the arm-by-time-since-launch term again measured any change in slope at launch.

#### Patient and public involvement statement

The study was designed and implemented with the Ivorian Ministry of Health at national, district and local levels, which approved implementation, helped select sites and supported community mobilisation and CHW recruitment. Patients were not involved in setting the research question or outcome measures.

## RESULTS

Figure 1 shows monthly facility-based under-five consultations in Madinani and in the 20 matched comparison districts over the pre-intervention period. Table 1 presents summary statistics by arm and period. Tables 2 and 3 present the estimated effects and the rank analysis.

**Figure 1.**
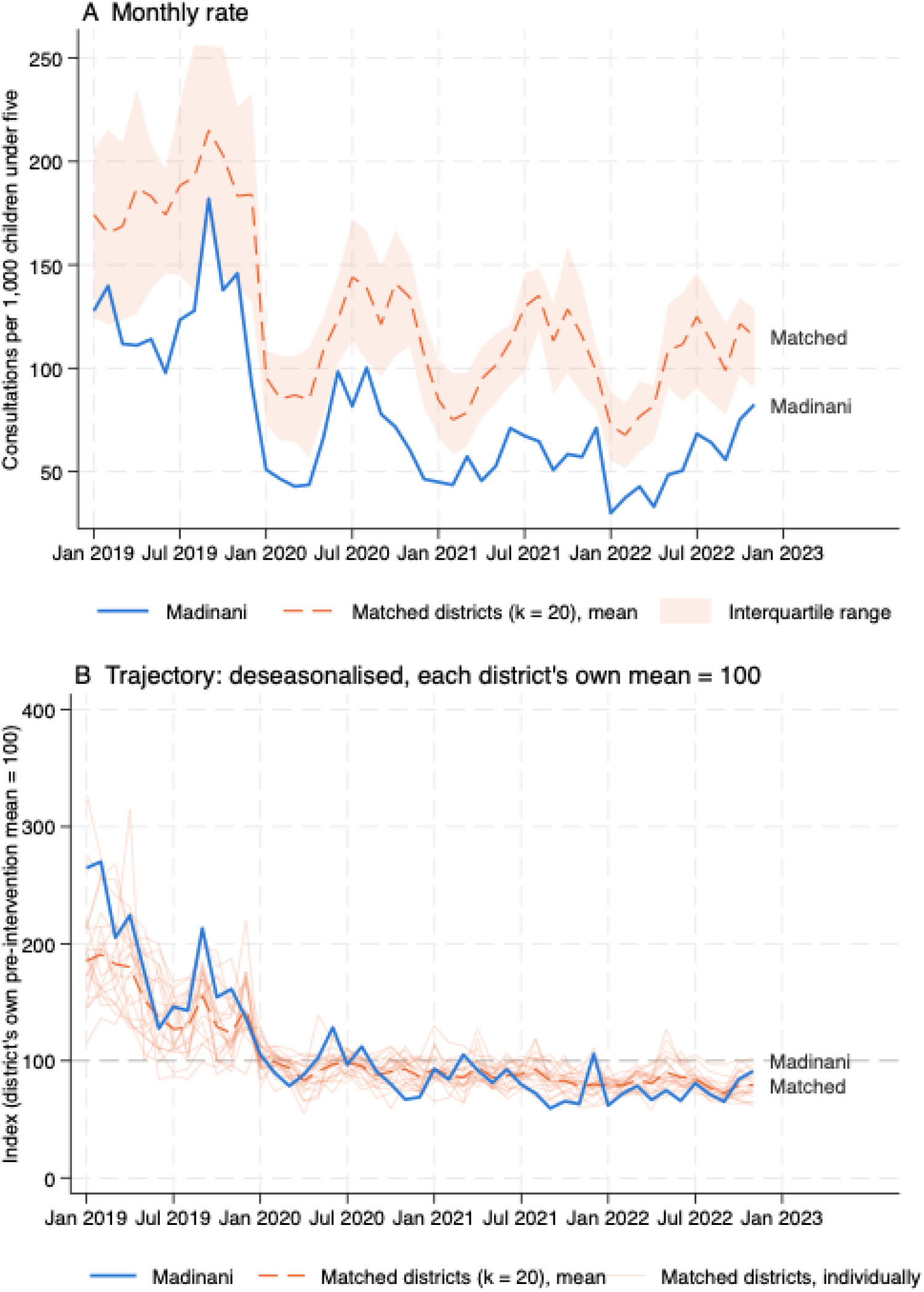
Pre-intervention trend in facility-based under-five consultations, Madinani and the 20 matched comparison districts, January 2019 to November 2022. (A) Monthly consultations per 1,000 children under five; shaded band, interquartile range of the matched districts. (B) Deseasonalised rate indexed to each district’s own pre-intervention mean (= 100); thin lines, individual matched districts.

Before ProCCM the mean monthly number of all-cause under-five consultations was 75.9 per 1,000 under-fives (SD 35.0 across months) in Madinani and 126.6 per 1,000 (SD 33.0 across district means) in the comparison districts; afterwards Madinani rose to 114.9 (SD 33.2) while the comparison districts fell to 112.5 (SD 29.0). Madinani began well below its comparison districts and ended at a comparable level: districts were matched on the shape of the pre-intervention trajectory rather than its level, so a level difference is not unexpected and is accounted for by the district random intercepts. A similar pattern was seen for diarrhoea, pneumonia, general-population consultations, first-trimester antenatal care and new family planning users.

In contrast, summary statistics indicated that institutional deliveries, EPNC, and the number of pregnant women aged 15-49 years receiving three doses or more of intermittent preventive treatment with sulfadoxine-pyrimethamine seemed to decline between the pre- and post-intervention period in both Madinani and the control districts.

**Table 1.** Mean monthly rates (SD) of the assessed outcomes by study arm and period, Madinani and the 20 matched comparison districts. Rates are per 1,000 children under five except: per 1,000 under-five consultations (rows marked per under-five consultation), per 1,000 expected pregnancies (first-trimester antenatal visit, IPTp-SP), per 1,000 expected live births (institutional deliveries, postnatal consultation), per 1,000 women aged 15-49 (family planning) and per 1,000 population (general-population consultations). SD is across months for Madinani and across district means for the comparison districts.

|  | Madinani |  | Comparison districts (k=20) |  |
|---|---|---|---|---|
| Outcome | Before (SD) | After (SD) | Before(SD) | After (SD) |
|  | Child health care service utilisation and general curative consultations, facility level |  |  |  |
| Under-five consultations, all causes | 75.9 (35.0) | 114.9 (33.2) | 126.6 (33.0) | 112.5 (29.0) |
| Curative consultations, general population | 52.4 (20.9) | 75.5 (16.4) | 69.9 (19.4) | 54.6 (14.0) |
| Confirmed malaria treated with ACT (per under-five consultation) | 718.0 (106.8) | 505.0 (78.1) | 580.2 (108.5) | 635.7 (129.4) |
| Diarrhoea treated with ORS and zinc (per under-five consultation) | 54.7 (32.7) | 45.9 (20.3) | 39.3 (22.3) | 34.4 (20.5) |
| Pneumonia treated with antibiotics (per under-five consultation) | 152.9 (46.6) | 129.7 (24.4) | 112.2 (28.9) | 86.6 (38.6) |
| Confirmed malaria treated with ACT (per under-five population) | 55.2 (28.7) | 58.6 (21.0) | 71.7 (20.8) | 71.9 (24.0) |
| Diarrhoea treated with ORS and zinc (per under-five population) | 3.6 (1.6) | 4.8 (1.6) | 4.5 (2.5) | 3.6 (2.1) |
| Pneumonia treated with antibiotics (per under-five population) | 11.2 (5.5) | 15.0 (5.8) | 14.4 (4.6) | 9.5 (4.6) |
|  | Child health care service utilisation, community level |  |  |  |
| Malaria treated with ACT | 0.1 (0.3) | 19.0 (7.8) | 1.3 (1.0) | 1.8 (1.3) |
| Diarrhoea treated with ORS and zinc | 0.9 (0.8) | 14.6 (6.4) | 1.5 (1.1) | 1.3 (0.8) |
| Pneumonia treated with antibiotics | 1.5 (1.5) | 43.7 (15.3) | 6.7 (4.4) | 13.6 (10.5) |
|  | Child health care service utilisation, facility and community level combined |  |  |  |
| Malaria treated with ACT | 54.5 (28.5) | 77.5 (24.9) | 70.5 (19.4) | 73.8 (24.6) |
| Diarrhoea treated with ORS and zinc | 4.5 (2.0) | 19.4 (6.7) | 6.0 (3.5) | 4.9 (2.6) |
| Pneumonia treated with antibiotics | 12.7 (5.3) | 58.8 (19.7) | 20.3 (6.3) | 23.1 (12.8) |
|  | Maternal health care service utilisation |  |  |  |
| First antenatal visit in the first trimester | 407.9 (97.5) | 450.8 (70.5) | 528.6 (194.4) | 315.8 (109.3) |
| Third dose of IPTp-SP | 607.0 (176.5) | 502.3 (94.9) | 805.6 (174.0) | 525.4 (123.8) |
| Institutional deliveries | 1048.8 (371.8) | 877.4 (133.0) | 1274.5 (263.0) | 1100.1 (181.7) |
| Postnatal consultation within 72 hours | 1120.9 (388.2) | 997.0 (147.3) | 1305.9 (272.4) | 1138.2 (202.5) |
| New users of family planning | 5.7 (5.1) | 7.9 (2.0) | 7.8 (2.1) | 6.1 (1.8) |

### ProCCM effects on child healthcare service utilization

At the facility level, all-cause under-five consultations in Madinani rose immediately after launch relative to the matched comparison districts (RRR 2.00, 95% CI 1.55 to 2.57, p<0.001). No evidence of further month-on-month change (RRR per month 1.007, 95% CI 0.992 to 1.023, p=0.36) was observed. Combining the two, utilisation at 24 months was 2.37 times (95% CI 1.72 to 3.27) what it would have been without the intervention (Supplementary Table S14 and Supplementary Figure S7). Under the common-slope specification the step change was smaller but still large (RRR 1.71, 95% CI 1.37 to 2.13, p<0.001) (Supplementary Table S14).

Treatment per 1,000 children under five increased for diarrhoea with ORS and zinc (RRR 1.91, 95% CI 1.21 to 3.03, p=0.006) and pneumonia with antibiotics (RRR 1.55, 95% CI 1.01 to 2.36, p=0.044), with no evidence of a step change for malaria treated with ACT (RRR 1.25, 95% CI 0.87 to 1.80, p=0.24), although its trend rose after launch (RRR per month 1.025, 95% CI 1.002 to 1.048), a rise that rests on extrapolating Madinani’s steep pre-intervention decline. As a share of under-five consultations, malaria fell substantially (RRR 0.63, 95% CI 0.48 to 0.83, p=0.001): the number treated did not rise at launch, but the proportion of consultations for malaria fell because consultations for other causes rose around it. Facility consultations did not fall when community care was introduced, so any shift of care was more than offset by demand generated by user fee removal and facility strengthening (Supplementary Figure S8).

The district-level ranking gives a concordant picture (Table 3). Madinani’s post:pre ratio for all-cause under-five consultations was 1.51, fifth highest of 102 districts (one-sided empirical p=0.049). Madinani ranked third for pneumonia and 19th for diarrhoea per 1,000 under-fives, but 62nd for malaria, matching the absence of a facility-level effect, and last of 102 for malaria as a share of under-five consultations.

Community-based curative care was close to absent in Madinani before ProCCM (Supplementary Figure S10). The mean monthly number of children treated by community health workers rose from 0.1 to 19.0 per 1,000 for malaria, 0.9 to 14.6 for diarrhoea and 1.5 to 43.7 for pneumonia (Table 1). In contrast, some community treatment of malaria was recorded in control districts in 77% of their pre-intervention district-months, none was without it throughout, and their community treatment of pneumonia rose from 6.7 to 13.6 per 1,000. The contrast is therefore one of scale rather than of presence and absence.

Because Madinani’s pre-intervention values are close to zero the corresponding ratios of rate ratios are very large and imprecise; for malaria the point estimate exceeds 100 000 and is not reported. Madinani had the highest post:pre ratio of all districts with usable community series for each condition (Table 3; Supplementary Figure S10).

Taking facility and community care together, the quantity of primary policy interest since it measures whether more sick children were reached overall, utilisation increased for all three conditions (Supplementary Figure S9): malaria treated with ACT (RRR 1.87, 95% CI 1.32 to 2.66, p<0.001), diarrhoea treated with ORS and zinc (RRR 8.37, 95% CI 5.29 to 13.24, p<0.001) and pneumonia treated with antibiotics (RRR 4.97, 95% CI 3.34 to 7.41, p<0.001). Under the common-pre-trend specification diarrhoea (RRR 7.09, 95% CI 4.79 to 10.51) and pneumonia (RRR 4.59, 95% CI 3.25 to 6.49) were little changed, but malaria fell to 1.31 (95% CI 0.96 to 1.79, p=0.088) and no longer reached conventional significance; the combined malaria result is the least secure of the three, consistent with the facility-level estimate and with a rank of 20th of 99 (p=0.20). On the ranking (Table 3) Madinani was first of 99 for pneumonia (p=0.010) and second for diarrhoea (p=0.020).

### Reproductive and maternal healthcare service utilization

The pre-intervention trend test (Supplementary Table S18) was satisfied for the third dose of IPTp-SP (trend ratio 1.000, p=0.90), institutional deliveries (0.996, p=0.21), early postnatal care (0.997, p=0.45) and new users of family planning (1.005, p=0.21). It was not satisfied for first-trimester antenatal care (1.015, 95% CI 1.008 to 1.023, p<0.001), for which both specifications are reported side by side below.

The evidence on early antenatal care depends on how the pre-intervention trend is handled. In the primary specification first-trimester visits rose by 40%, but the interval included no effect (RRR 1.40, 95% CI 0.99 to 1.99, p=0.061) and waned after launch (RRR per month 0.974, 95% CI 0.954 to 0.995); under the common-slope specification the estimate was a doubling and precisely estimated (RRR 2.00, 95% CI 1.48 to 2.71, p<0.001). This is the one outcome for which the violation materially changes the conclusion, and it acts opposite to the primary outcome: Madinani’s pre-intervention antenatal trend rose faster than its comparators’, so projecting it forward raises the counterfactual and shrinks the effect (Supplementary Figure S11). Madinani ranked eighth of 102 (p=0.078). We report the primary estimate and regard this effect as suggestive rather than established.

ProCCM was associated with an increase in third-dose IPTp-SP (RRR 1.36, 95% CI 1.06 to 1.75, p=0.017), unchanged under the alternative specification and with parallel trends satisfied, although Madinani ranked only 37th of 102 (p=0.36) because the absolute rate fell in both arms. There was no evidence of an effect on institutional deliveries (RRR 1.01, 95% CI 0.76 to 1.35, p=0.93; rank 84) or postnatal consultation within 72 hours (RRR 1.03, 95% CI 0.77 to 1.38, p=0.84; rank 70). New family planning users increased (RRR 1.94, 95% CI 1.26 to 2.99, p=0.003; rank 7), rising from 5.7 to 7.9 per 1,000 women aged 15-49 while falling from 7.8 to 6.1 in the comparison districts (Supplementary Figure S11).

### General population healthcare utilization

ProCCM was associated with a substantial increase of curative consultations in the general population (RRR 2.20, 95% CI 1.76 to 2.74, p<0.001), with no evidence of further month-on-month change (RRR per month 1.003, p=0.71) (Table 2) and a combined effect at 24 months of 2.34 (95% CI 1.76 to 3.11) (Supplementary Figure S12 and Supplementary Table S14). Under the common-slope specification the estimate was 1.92 (95% CI 1.58 to 2.32) (Supplementary Table S14). Madinani also had the highest post:pre ratio of all 102 districts for this outcome (Table 3; p=0.010).

**Table 2.**
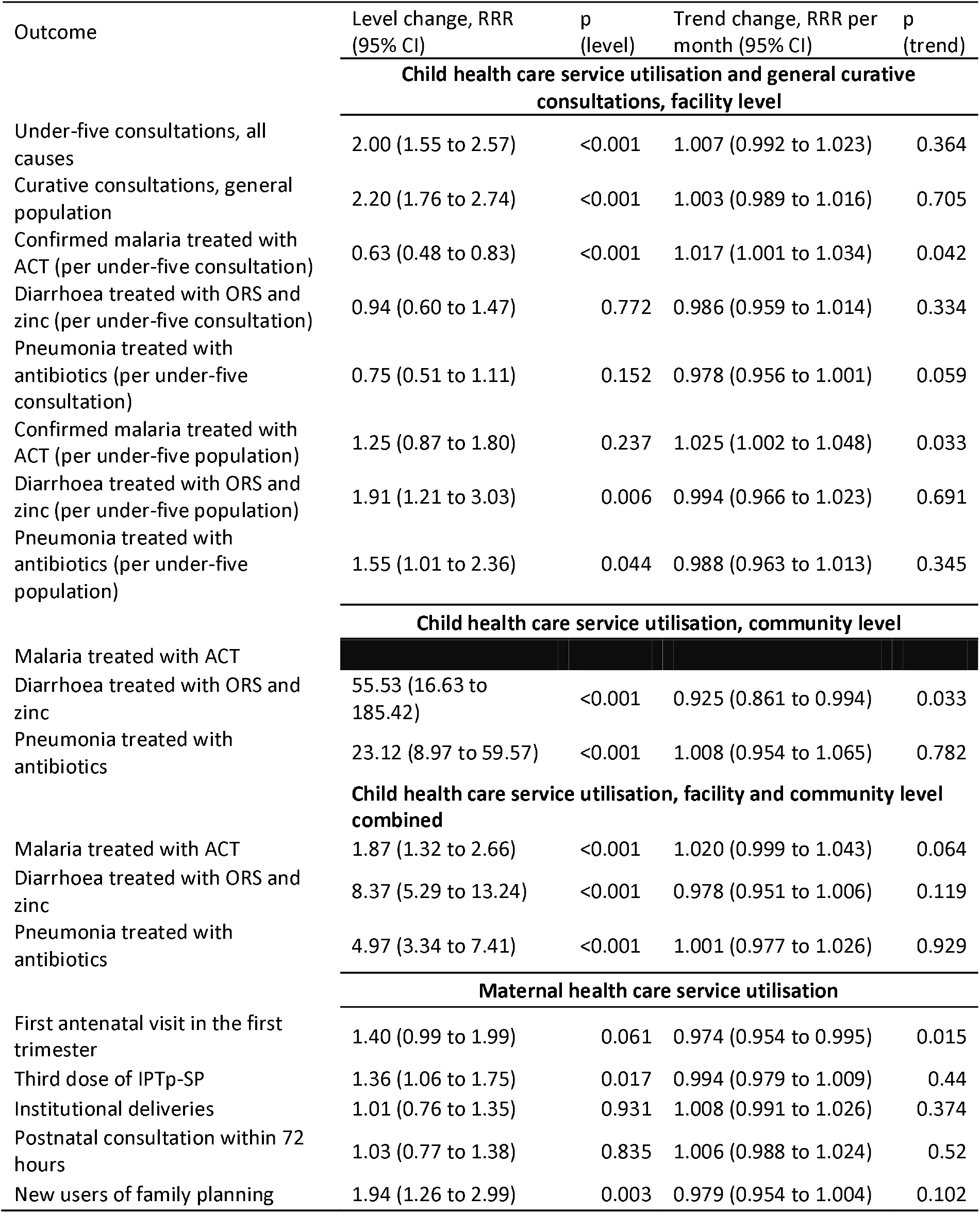
Estimated effects of ProCCM on healthcare utilisation in Madinani relative to the 20 matched comparison districts: level change and change in trend, as ratios of rate ratios (primary specification, pooled over 20 imputed datasets). Community-level malaria is not shown: Madinani’s pre-intervention count was close to zero and the estimate is not interpretable.

**Table 3.**
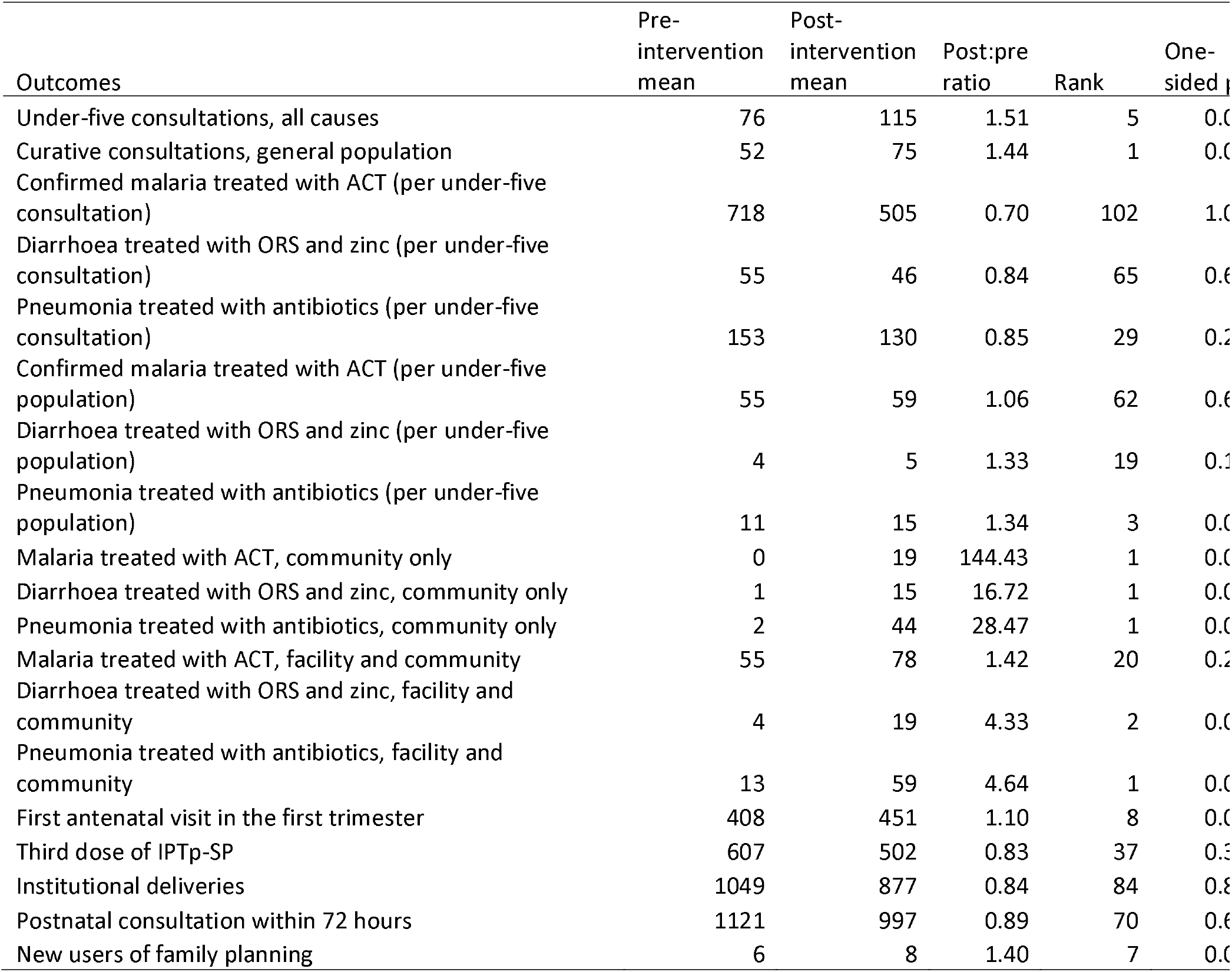
Madinani’s rank among all eligible districts on the ratio of post-to pre-intervention mean monthly rates (rank 1=largest relative increase; 102 districts for facility-based outcomes, 99 for community and combined outcomes). One-sided empirical p=rank/number of districts.

### Sensitivity analysis results

Six sensitivity analyses were performed (Figure 2; Supplementary Tables S13 and S17; Supplementary Figures S15 and S16). First, varying the comparison group gave 2.03 with 5 districts, 2.00 with 10 and 20, 2.01 with 15, 1.98 with 30, 2.14 with 50 and 2.29 with all 101 unmatched, so the result does not depend on how many comparison districts are used and matching makes the estimate more conservative.

Second, selecting comparison districts on level and slope rather than trajectory gave 1.84 (95% CI 1.41 to 2.39), with 13 of 20 districts chosen by both rules. Third, re-selecting per outcome did not change the direction of any estimate (Supplementary Table S13). Fourth, on the dataset restricted to consistently reporting facilities the two agreed for the primary outcome (2.02 against 2.00), while other outcomes were systematically somewhat larger, the largest discrepancy being 1.6 standard errors (Supplementary Figure S16). Fifth, the two trend specifications agreed for the primary outcome (2.00 against 1.71; Supplementary Figure S15); outcomes where they differ are identified above.

Sixth, giving every district its own slope changed no conclusion (Supplementary Table S17). District slopes varied (likelihood ratio test p<0.001), and for the primary outcome Madinani’s lay 1.7 standard deviations below the comparison average, the third steepest of 21. The level change was 2.01 (95% CI 1.61 to 2.52) with Madinani’s slope estimated separately and 1.94 (1.56 to 2.42) with it partially pooled, with no evidence of a change in slope at launch (RRR per month 1.008, 95% CI 0.994 to 1.021). Across the 11 outcomes examined, level changes were within 8% of the primary estimates. With partial pooling, first-trimester antenatal care was 1.50 (1.12 to 2.01) and combined malaria care 1.83 (1.34 to 2.49), both close to their primary estimates. These models were fitted to a single dataset rather than to the 20 imputed datasets.

**Figure 2.**
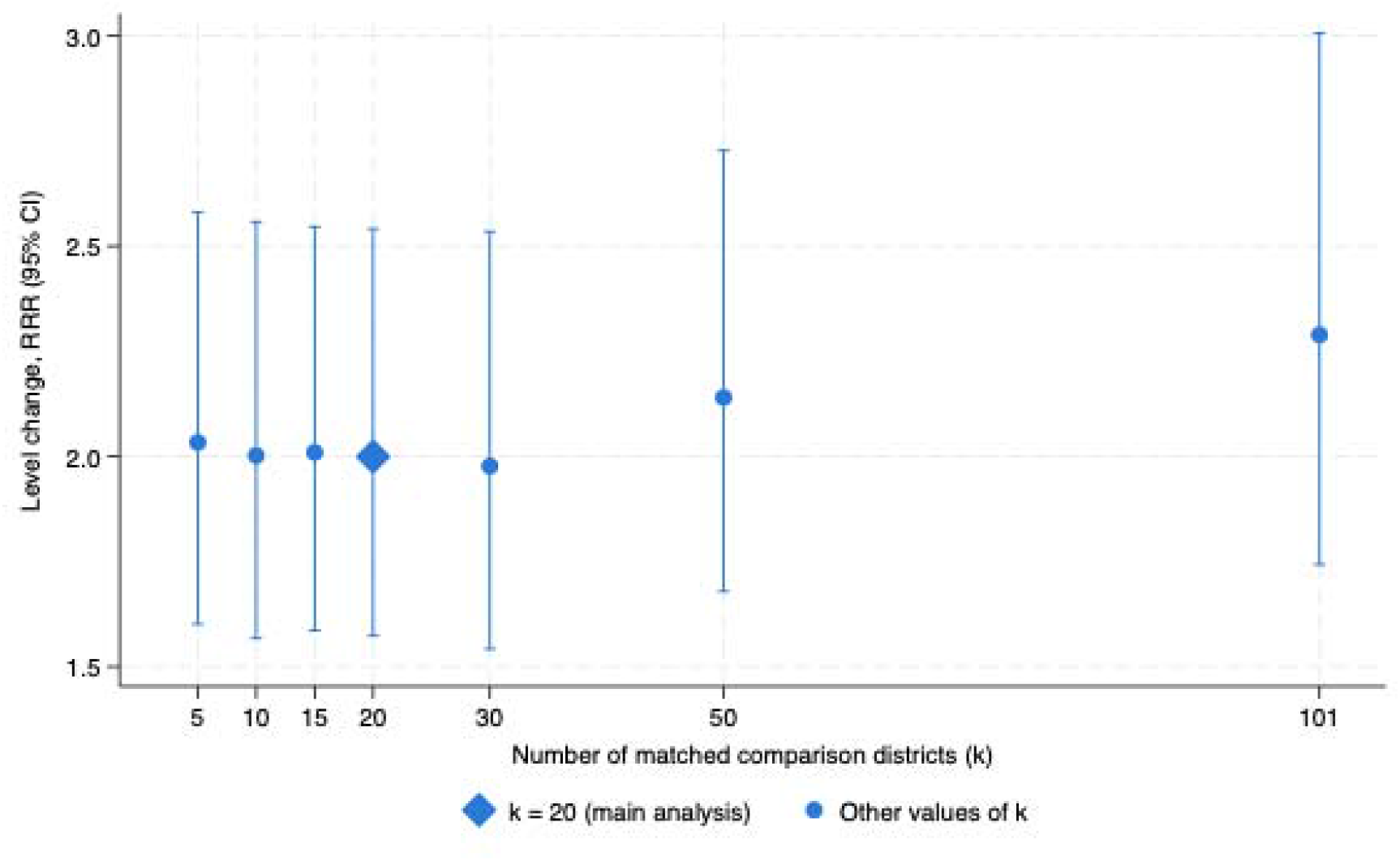
Level change in facility-based under-five consultations (ratio of rate ratios, 95% CI) by number of matched comparison districts (k), from 5 to all 101 eligible districts.

## DISCUSSION

In Madinani, ProCCM was followed by a doubling of facility-based under-five consultations relative to 20 matched comparison districts (RRR 2.00, 95% CI 1.55 to 2.57), appearing as a step at launch and sustained without further growth. Treatment for diarrhoea and pneumonia increased at facility level, and all three conditions increased substantially when facility and community care are counted together. General-population consultations more than doubled (RRR 2.20, 95% CI 1.76 to 2.74) and third-dose IPTp increased modestly (RRR 1.36, 95% CI 1.06 to 1.75). The effect on first-trimester antenatal care was sensitive to the trend specification, and we regard it as suggestive. There was no evidence of an effect on institutional deliveries, postnatal care, or the number of children treated for malaria at facilities, although malaria fell sharply as a share of under-five consultations.

Our findings on child healthcare utilisation complement those from Mali (20, 22). Pre-post studies of the urban Mali rollout documented falls in under-five mortality and a rise in early effective antimalarial treatment (20, 21). The present study adds a quasi-experimental analysis against historical and geographic controls.

The very large ratios for community-based treatment follow from the near-absence of these services beforehand: they indicate a service brought to scale rather than one multiplied and are best read with the absolute numbers in Table 1. The more informative question is what happened to facility care once community treatment became available. It could in principle have reduced facility attendance, yet facility consultations rose, so the combined increase reflects wider coverage rather than a transfer between levels. The same logic explains the malaria results: the number treated at facilities did not change (RRR 1.25) while combined care rose (RRR 1.87), and malaria fell as a share of consultations (RRR 0.63) because other consultations rose around it. The model-free ranking provides a check independent of the trend specification, and for the primary outcome the two agree: fifth of 102 (p=0.049) against a CITS estimate of 2.00.

Proactive case detection through home visits, doorstep care and dedicated supervision are the components most likely to have improved access. Payment and incentives play a substantial part in CHW motivation and performance (33, 34), and proactive case detection has documented effects on effective access to child healthcare (35).

User fee removal has been documented as effective in improving child healthcare utilisation at facility level: in Burkina Faso the free healthcare policy increased use of child healthcare services at facility level (36–38). However, our findings on the non-effect of ProCCM for treating malaria at the facility level, alongside the increase seen when facility and community care are counted together (RRR 1.87), highlight the effectiveness of ProCHWs in early detection and treatment of malaria in the community.

Regarding the ProCCM effect on reproductive and maternal health, the direction of the estimated effect on the proportion of pregnant women initiating antenatal care in the first trimester, though sensitive to how the pre-intervention trend is handled, is consistent with those obtained in the Malian setting (39), where ProCCM was associated with increases in any antenatal care and in four or more visits. The proactivity component is the most likely route by which early antenatal attendance would be raised.

The lack of a ProCCM effect on other reproductive and maternal health outcomes, such as institutional deliveries and postnatal care, may be due to two factors: pre-existing user fee removal for maternal care, and persistent structural barriers. Fees for prenatal care and facility births were removed nationally in 2011 (40), which may partly explain the absence of a further difference after ProCCM launch (41, 42). Before ProCCM fees were removed only for uncomplicated childbirth; ProCCM extended this to complications of pregnancy and childbirth, which the indicators used here do not capture, an important limitation. Structural barriers also persist: an audit of perinatal mortality at the site identified social norms and women’s social position, alongside distance and transport difficulties in remote villages, as continuing obstacles to institutional delivery. These findings have informed a re-adaptation of ProCCM to be assessed in the next planned evaluation.

The increase in outpatient utilisation for all ages and causes is an important finding, showing value beyond children and women of reproductive age. Removing user fees addresses financial barriers, while infrastructure and provider capacity components would improve the quality of care, and a positive perception of quality is associated with greater use of outpatient services (43), including in the context of ProCCM intervention delivery (22).

### Strengths and methodological considerations

The study has several strengths. It used a quasi-experimental design over 47 pre-intervention and 24 post-intervention months, long enough to characterise levels, trends and change points. Comparison districts were selected by a pre-specified, outcome-blind rule rather than judgement, and the estimate was stable as the group varied from 5 districts to all 101. Contextual changes were monitored, and no concurrent intervention was found at the site. Alongside the models, we report a model-free ranking of all eligible districts, letting readers judge the effect without relying on the time-series specification. Missing and extreme values were multiply imputed rather than discarded, with estimates pooled across 20 datasets. The study also demonstrates the use of Côte d’Ivoire’s DHIS-2 for evaluating public health interventions.

Several limitations should be noted. First, Madinani’s facility-based under-five consultation rate was falling faster before the intervention than almost any other district, and matching narrowed but did not eliminate that difference; parallel trends were violated for 9 of 19 outcomes. Because the primary model projects each arm’s slope forward, this moves estimates away from the null where Madinani declined faster and towards it where Madinani rose faster. Every outcome is therefore reported under a second specification with a common slope; for the primary outcome the two agree (2.00 against 1.71), while for first-trimester antenatal care and combined malaria they do not, and those results should be read as the range between them. Residual confounding from whatever caused Madinani’s earlier decline cannot be excluded. Second, the analysis rests on routine facility data, subject to completeness and consistency problems (44); repeating it on consistently reporting facilities gave systematically somewhat larger estimates, so the primary analysis is the more conservative. Coverage by those facilities was itself uneven: the median district had 91% of its reported volume in consistently reporting facilities, but in the least well covered district this fell to 29%. Third, the design does not involve random assignment, so unmeasured differences could contribute; this was mitigated by comparison with the district’s own history, a matched group and every other district. Fourth, strengthened supervision and reporting may have improved community recording relative to comparison sites, so part of the community-level increase may reflect more complete reporting; this applies least to the facility-based outcomes, reported through an established system throughout, and most to the community-only outcomes, which we report descriptively. Finally, every intervention-specific coefficient is identified from 71 monthly observations of one district, and the random intercept does not account for within-series correlation, so intervals around the step change may be too narrow; the ranking, free of that assumption, gives a concordant but more conservative answer.

## CONCLUSION

This study provides controlled evidence on an intervention that removed geographical and financial barriers to child, maternal and general healthcare. By combining user fee removal, professional community health workers and upgraded primary care, ProCCM increased access to child healthcare and to general healthcare across all ages, with a less certain effect on early antenatal care. These findings could inform Ivorian policymakers considering adaptation of evidence-based community health interventions.

## Data Availability

Data are available upon reasonable request. Aggregated DHIS-2 data can be obtained from the corresponding author with the approval of the Director of the Health Information System of the Ivorian Ministry of Health.

## Abbreviation

ACT: Artemisinin-based Combination therapy
CI: Confidence Interval
CITS: Controlled Interrupted Time Series
CHW: Community Health Worker
DHIS-2: District Health Information System
EPNC: Early Postnatal Care
ICCM: Integrated Community Case Management
IPTp-SP: Intermittent Preventive Treatment in pregnancy with Sulfadoxine-Pyrimethamine
MoHPHUHC: the Ministry of Health, Public Hygiene, and Universal Health Coverage
ORS: Oral Rehydration Solution
ProCCM: Proactive Community Case Management
ProCHW: Professional Community Health Worker
RMNCH: Reproductive, Maternal, Neonatal, and Child Health
RRR: Ratio of rate ratios
SD: Standard Deviation
SSA: Sub-Saharan African
U5M: Under-Five Mortality
UNICEF: United Nations Children’s Fund WHO World Health Organization

## Contributions

ABS, JAKK, SC, MAAT, and AJ conceptualized the study. LG and BT performed data management. ABS led data analyses with oversight from SC. ABS wrote the first draft of the manuscript with inputs and critical reviews from BT, AFO, LG, TS, ABC, YK, CR, JAKK, RY, SM, JB, ET, SC, AJ, and MAAT. All co-authors approved the manuscript. ABS is the guarantor.

## Competing interests

ABS, BT, LG, ABC, YK, CR, JAKK, JB, TS, MAAT, and AJ were employed by Muso during the investigation period. All other authors have no competing interests to declare.

## Ethics approval

This study involves human participants and was approved by the Comité National d’Ethique des Sciences de la Vie et de la Santé (CNESVS), Côte d’Ivoire, reference number 168-22/MSHPCMU/CNESVS-km. This analysis used aggregated, anonymized routine data; no individual-level data were accessed, so individual informed consent was not required.

## Data availability statement

Data are available upon reasonable request. Aggregated DHIS-2 data can be obtained from the corresponding author with the approval of the Director of the Health Information System of the Ivorian Ministry of Health. The Stata code is available from the corresponding author.

## Funding

This study did not receive any specific funding. However, Muso Health receives unrestricted donations from more than 50 foundations and individual partners to support its research, service delivery, and technical assistance activities.

## Acknowledgments

We thank the ProCHWs at the MUSO Côte d’Ivoire site and their supervisors for participating and contributing to study implementation. We also thank the Muso Program, Innovation and Training, and Technical Assistance teams for their operational and logistical support throughout the study period. We also thank district health information officers, who report facility data to DHIS-2, and Noe Yecoula and Ben Ally Sadyhat Bamba from the Division of Health Information Systems of MoHPHUHC, who facilitated data extraction and data review.

## Supplementary materials

### Supplementary Methods

#### SM1. Study setting

Madinani is located in the Denguélé region in the north-west of Côte d’Ivoire, approximately 700 kilometres from Abidjan, the country’s economic capital and largest city (Yamoussoukro is the political capital). The district’s population is estimated at 106 000 inhabitants (2021 census, unpublished) and is predominantly Muslim, with Malinké and Senoufo as the dominant ethnic groups. The district covers approximately 7 000 km², a population density of about 15 inhabitants per km².

Madinani health district was selected by Muso in collaboration with the MoHPHUHC on predefined programme implementation criteria: a high under-five mortality rate, low access to healthcare, absence of competing interventions, minimal security risk, and geographic diversity. The ProCCM intervention was launched in Madinani in December 2022.

At launch, Madinani had 25 health facilities: two district hospitals, 13 community health centres and 10 rural health posts; the 23 community health centres and rural health posts constitute the primary healthcare level. According to the Departmental Health Directorate, the distance to the nearest facility varies widely and in many settings remains high, with some villages nearly 80 kilometres from their nearest facility, making the journey impracticable in many contexts. This far exceeds the WHO recommendation that all populations be within a five-kilometre radius of their nearest primary healthcare facility. All 25 facilities are government-owned, and there is no formal private-sector provider reporting into DHIS-2 in Madinani, so the DHIS-2 data used here capture essentially all formal care delivered in the district.

During a preliminary situational assessment conducted before the launch of ProCCM, Muso and the MoHPHUHC observed that the recent construction of several primary healthcare centres was inadequate and that most facilities in Madinani required renovation, with several failing to meet national construction standards. Most of the centres visited were new, and many lacked essential equipment, notably diagnostic equipment. Not all primary healthcare centres had access to water and electricity; some state nurses had to seek connections to local utility networks personally, resulting in an intermittent supply at their facilities.

#### SM2. Data management: the outlier rule and the imputation model

Data management was carried out at the level of the individual facility-month, before any aggregation to district level, and addressed two problems: values that were missing, and values that were present but implausible.

Detection of extreme values. Extreme values were identified by a robust, within-facility rule on the log scale. Each facility’s series was deseasonalised and each observation compared with that facility’s own median, scaled by its median absolute deviation. A value was flagged only if all four of the following held: its robust z score exceeded 6; the facility contributed at least 12 months in the relevant period; the count itself was at least 10; and the value was also at least five times, or at most one fifth of, that facility’s own median.

The last condition matters because facility counts are heavy-tailed. In a small facility reporting 0, 1, 0, 2 the median absolute deviation is near zero, so a z-score rule alone flags ordinary variation. Requiring the value also to be extreme on an interpretable scale removed almost all such cases while retaining genuine errors; the rule flagged between 0.6% and 2.0% of facility-months, depending on the outcome. Medians and deviations were computed separately for the pre- and post-intervention periods in every district, not only in Madinani, so that the two arms were screened symmetrically and a genuine post-intervention increase in Madinani was not treated as an error and imputed away.

Imputation. Flagged extreme values were set to missing and, together with genuinely missing facility-months, imputed by predictive mean matching with five donors, separately within each district. The predictors were calendar month, calendar time, an indicator for the pre- or post-intervention period, the facility’s own mean of the variable being imputed, the facility’s overall caseload, and the facility’s reported completeness of monthly returns. Caseload was measured as the mean of the logarithm of monthly under-five consultations, that being the most consistently reported column in the extract and therefore available even for facilities whose other columns are sparse. The facility’s own mean and its caseload were both computed separately for the pre- and post-intervention periods, because a single value spanning both would pull post-launch gaps in the intervention district back towards its pre-launch level and bias the estimate towards the null.

Twenty imputed datasets were generated. Every model was fitted in each imputed dataset and the estimates combined by Rubin’s rules with Barnard-Rubin degrees of freedom, so that the reported confidence intervals include the uncertainty created by filling the gaps. The fraction of the total variance attributable to the imputation was small for every outcome (mean 0.03, maximum 0.10), and the pooled confidence intervals were on average 6% wider than those obtained from a single averaged dataset.

The two datasets. Two district-level datasets were constructed. The first uses every facility, with gaps and extreme values imputed as above, and is the basis of the primary analysis. The second retains only facilities reporting at least 90% of the months in their active window with no gap longer than three consecutive months, and is therefore imputed almost nowhere; the entire analysis was repeated on it as a completeness sensitivity analysis. Coverage by consistently reporting facilities was itself uneven: the median district had 91% of its reported volume in such facilities, but in the least well covered district this fell to 29%, and where coverage is low the restricted series is built from a minority of that district’s care.

#### SM3. The analysis model in full

The model contains a random intercept for each health district, so that every comparison district contributes its own monthly series and is not aggregated with the others; fixed effects for calendar month, capturing seasonality common to both arms; an indicator for study arm, with the comparison districts as the reference category; a linear term for calendar time and its interaction with study arm, so that the two arms may follow different pre-intervention trends; an indicator for the post-intervention period, entered both as a main effect and interacted with study arm, the interaction being the average step change in utilisation attributable to ProCCM; and a linear term for time elapsed since launch, likewise entered as a main effect and interacted with study arm, allowing the effect to grow or decay over the 24 post-intervention months.

Entering the post-intervention indicator and time since launch as main effects as well as interactions is what keeps any national change coinciding with January 2023 in the comparison arm, where it belongs, rather than in the intervention estimate. The second specification is identical except that the interaction between study arm and calendar time is omitted, so that both arms share a single pre-intervention slope.

Models were fitted with the menbreg command in Stata 19.5, with the logarithm of the denominator population as an offset and coefficients exponentiated as rate ratios; the exponentiated arm-by-period interaction, the estimated effect of ProCCM, is a ratio of rate ratios (RRR). Where a negative binomial model failed to converge, a linear mixed model of the log rate was to be fitted instead, for which exp(beta) estimates the same quantity; in the analyses reported here every model converged as a negative binomial and no fallback was required.

Because the level change and the trend change are not two separate findings - on the log scale the effect H months after launch is the level change plus H times the change in slope - we also report the combined effect 24 months after launch, obtained as a linear combination of the two coefficients so that its confidence interval accounts for their covariance rather than treating them as independent.

---

**Supplementary Figure 1.**
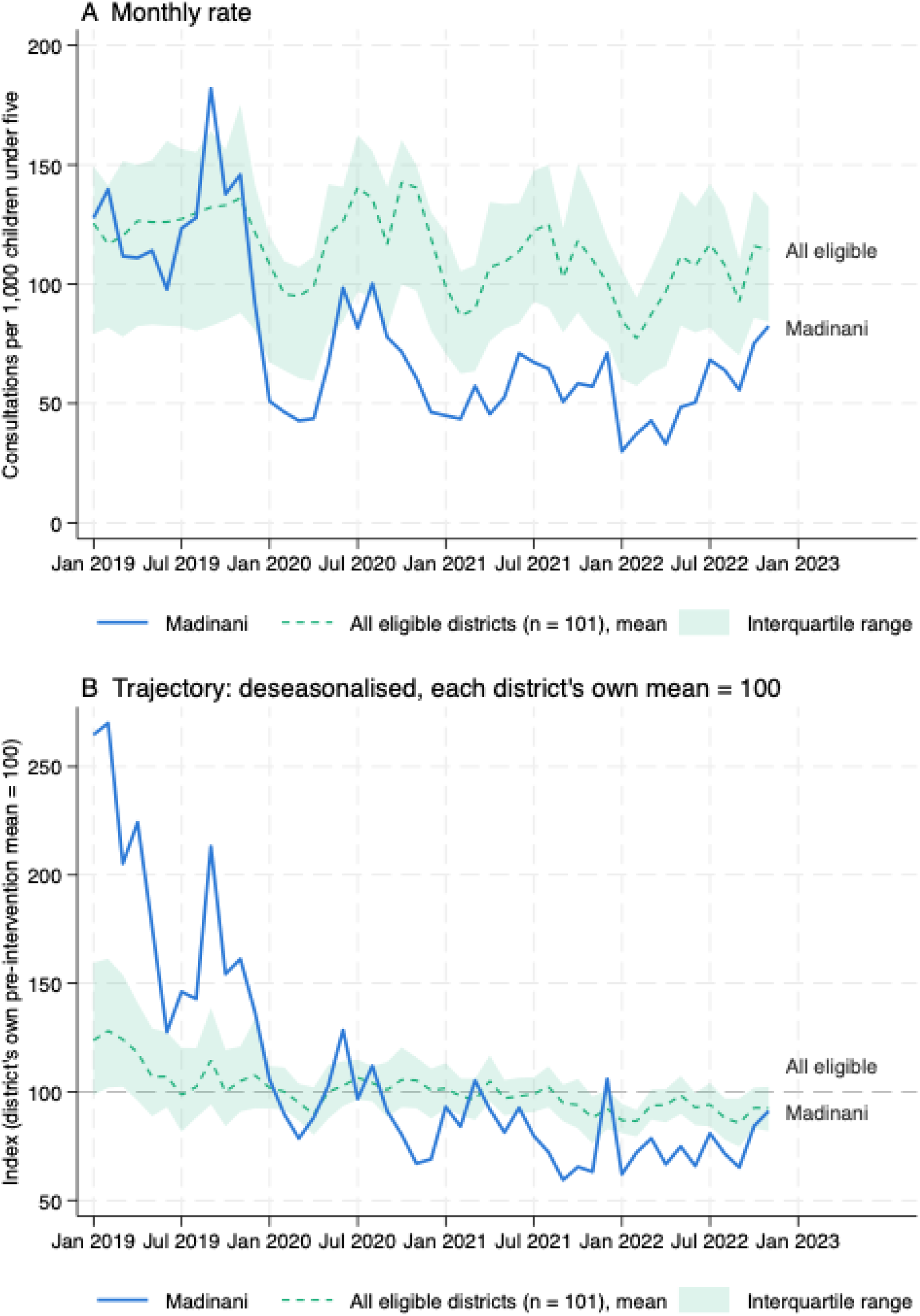
Pre-intervention trend in facility-based under-five consultations, Madinani and all 101 eligible districts, January 2019 to November 2022. (A) Monthly consultations per 1,000 children under five; shaded band, interquartile range of the eligible districts. (B) Deseasonalised rate indexed to each district’s own pre-intervention mean (= 100).

**Supplementary Figure 2.**
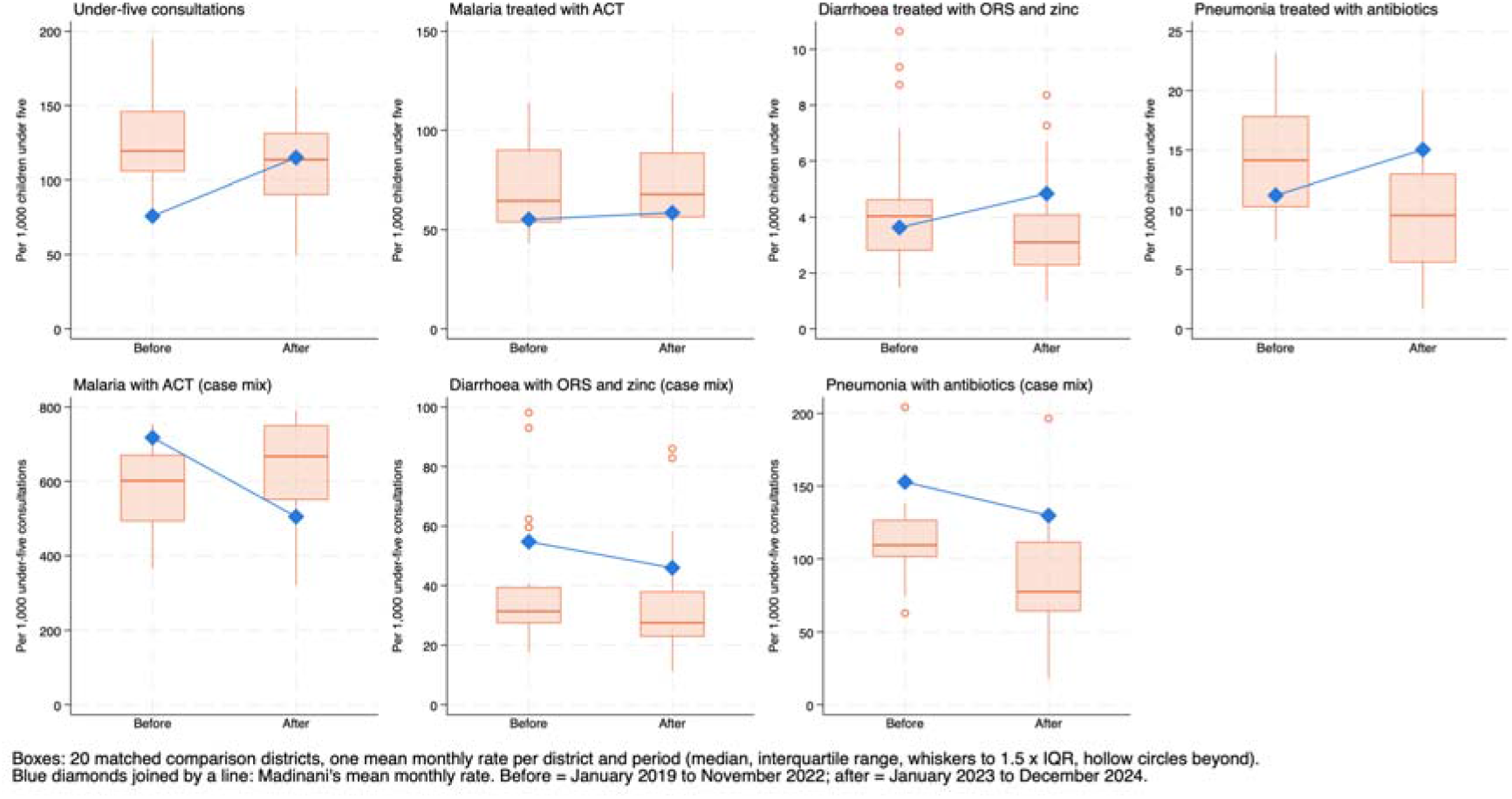
Mean monthly rates of child healthcare utilisation at facility level before (January 2019 to November 2022) and after (January 2023 to December 2024) launch. Boxes: the 20 matched comparison districts, one mean per district and period (median, interquartile range, whiskers to 1.5 × IQR). Diamonds: Madinani.

**Supplementary Figure 3.**
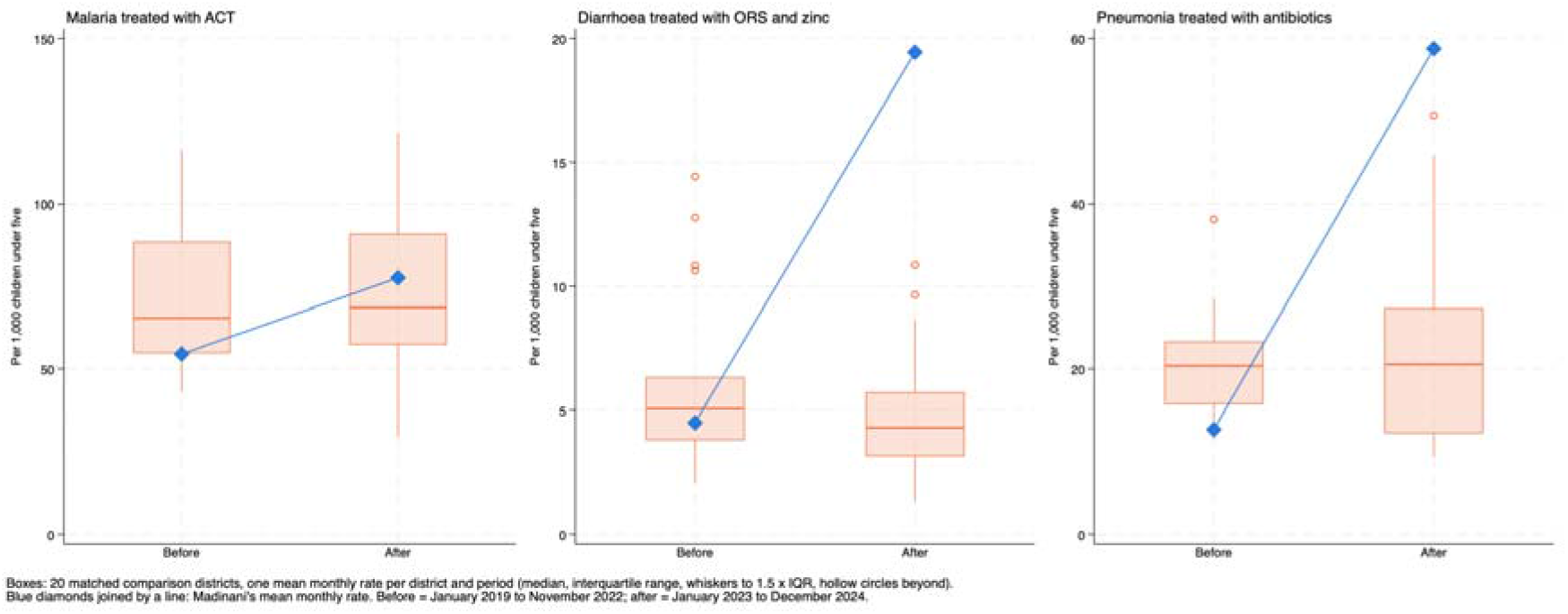
Mean monthly rates of child healthcare utilisation at facility plus community level before (January 2019 to November 2022) and after (January 2023 to December 2024) launch. Boxes: the 20 matched comparison districts, one mean per district and period (median, interquartile range, whiskers to 1.5 × IQR). Diamonds: Madinani.

**Supplementary Figure 4.**
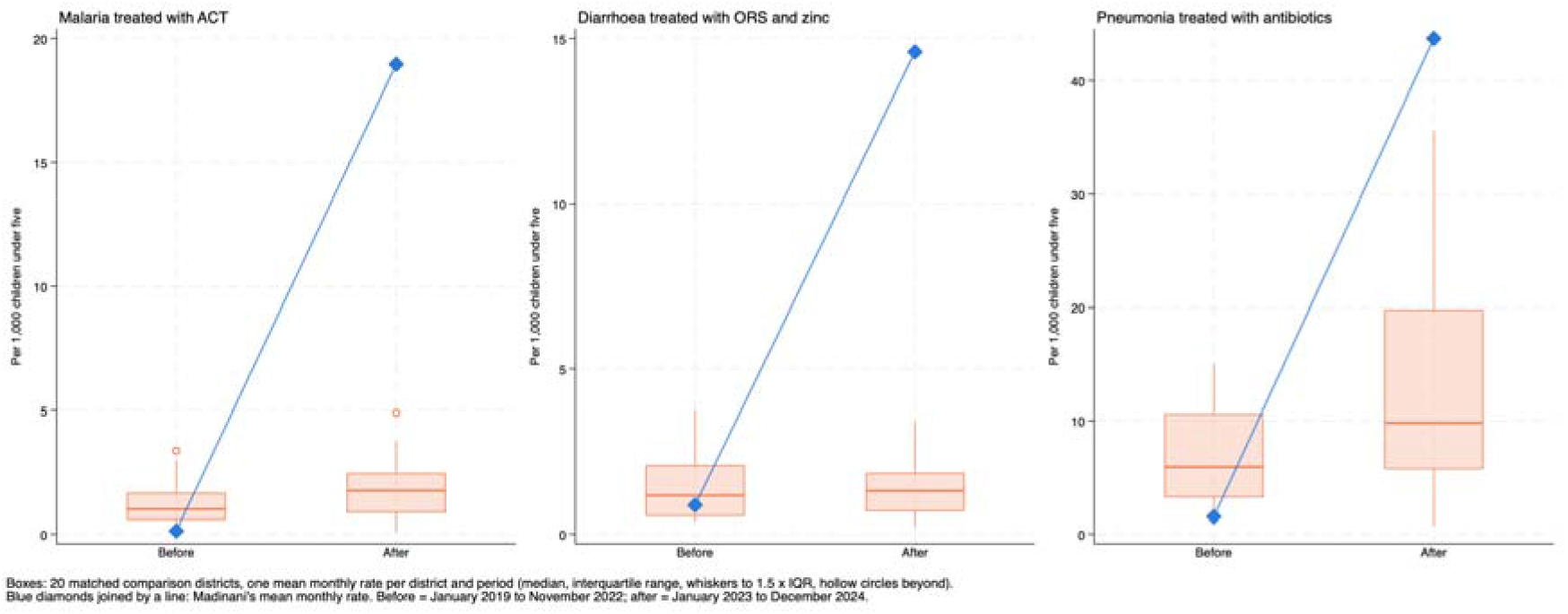
Mean monthly rates of child healthcare utilisation at community level only before (January 2019 to November 2022) and after (January 2023 to December 2024) launch. Boxes: the 20 matched comparison districts, one mean per district and period (median, interquartile range, whiskers to 1.5 × IQR). Diamonds: Madinani.

**Supplementary Figure 5.**
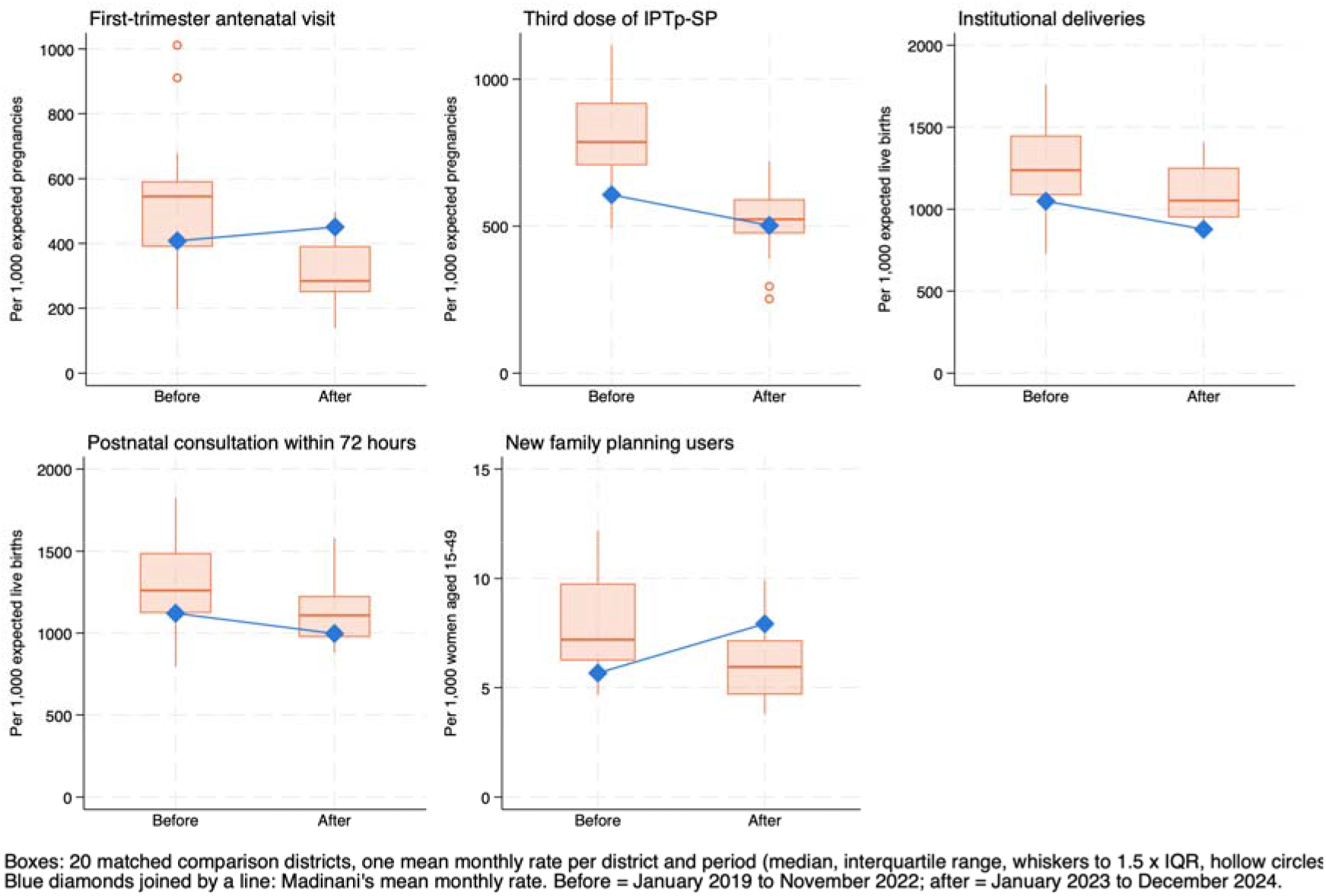
Mean monthly rates of maternal healthcare utilisation before (January 2019 to November 2022) and after (January 2023 to December 2024) launch. Boxes: the 20 matched comparison districts, one mean per district and period (median, interquartile range, whiskers to 1.5 × IQR). Diamonds: Madinani.

**Supplementary Figure 5.**
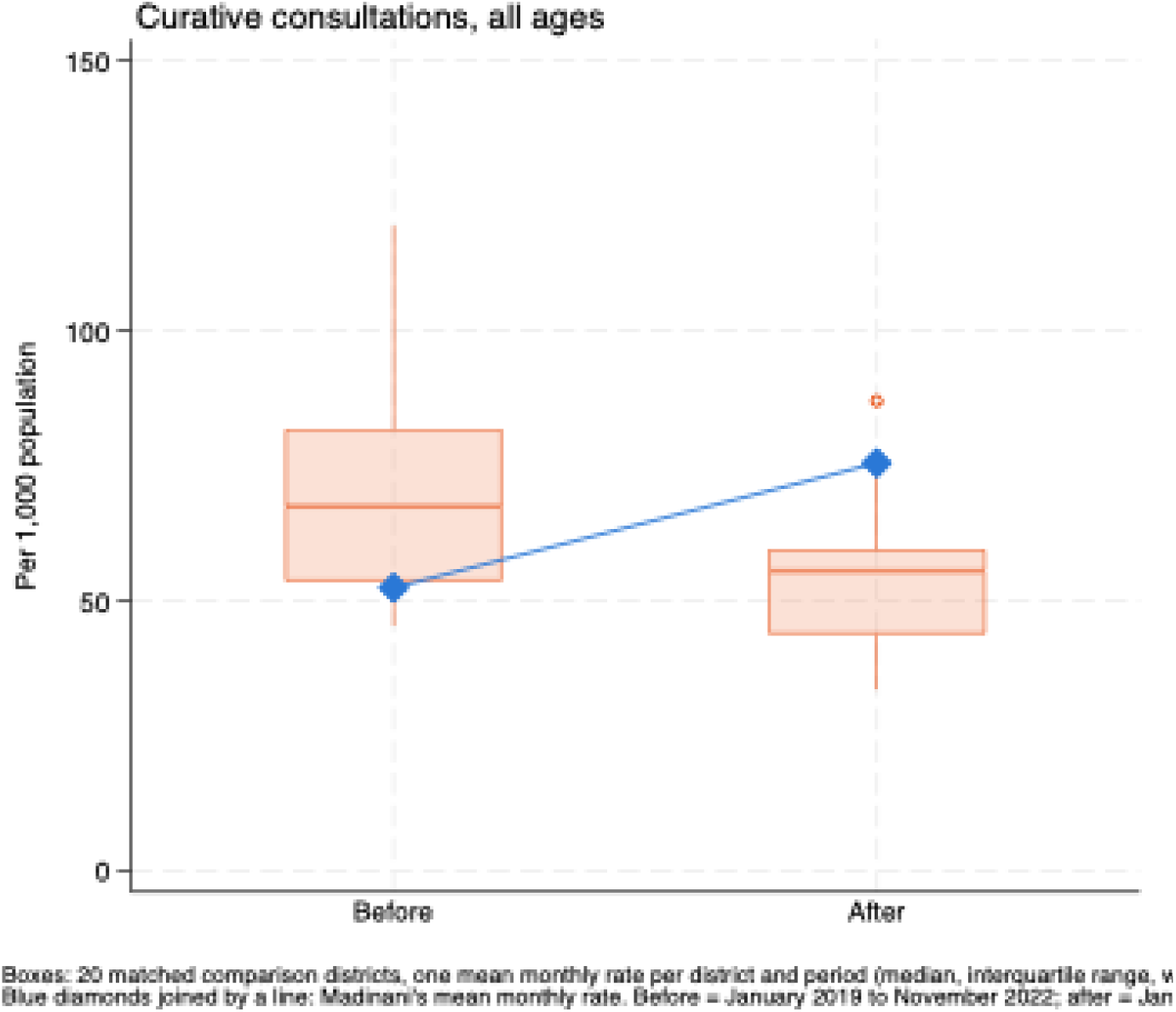
Mean monthly rates of curative consultations in the overall population before (January 2019 to November 2022) and after (January 2023 to December 2024) launch. Boxes: the 20 matched comparison districts, one mean per district and period (median, interquartile range, whiskers to 1.5 × IQR). Diamonds: Madinani.

**Supplementary Figure 7.**
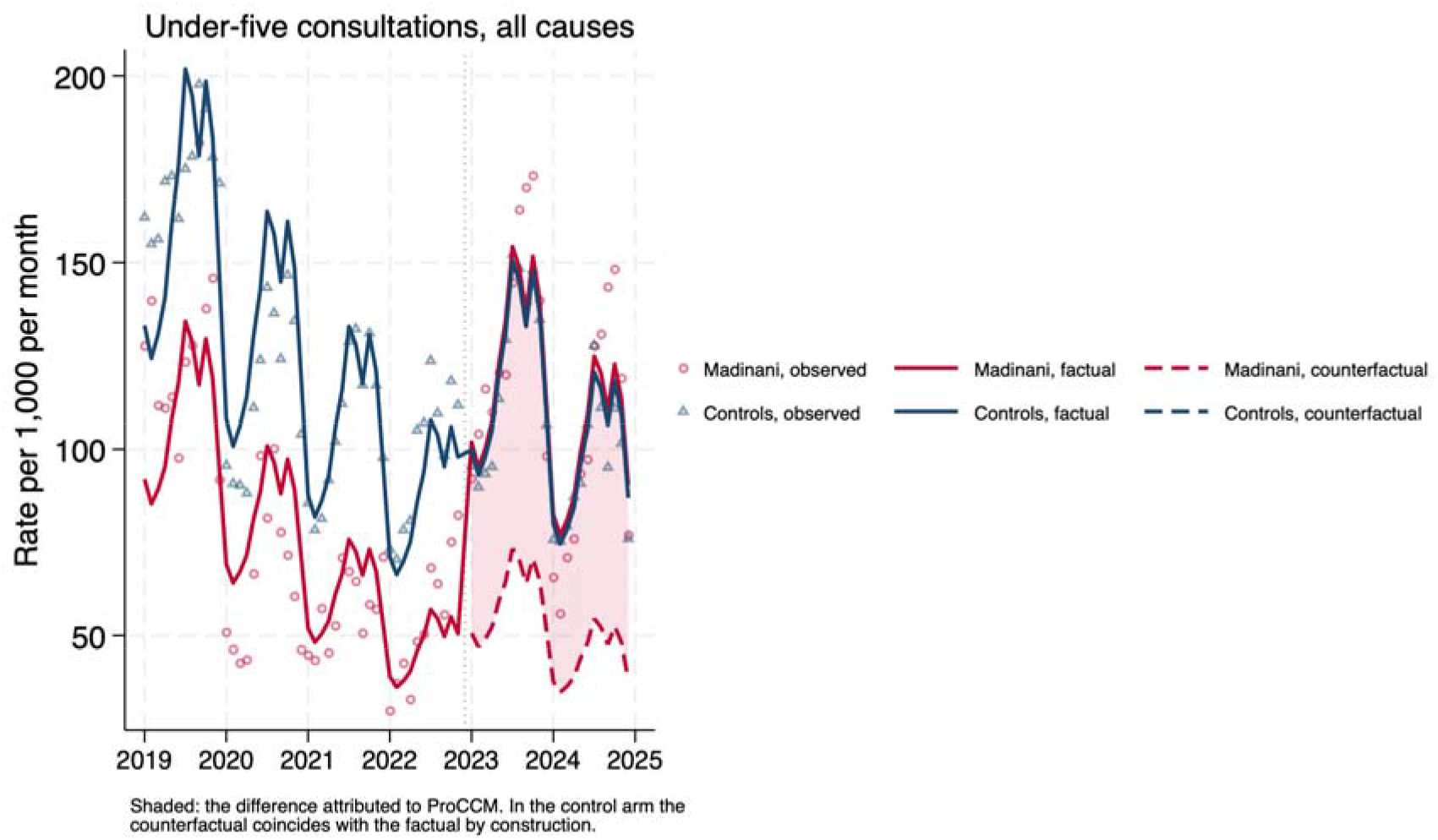
Monthly under-five consultations trend (Intervention versus control), from January 2019 to December 2024, with the ProCCM intervention implemented in December 2022 (vertical dashed line), created by authors.

**Supplementary Figure 8.**
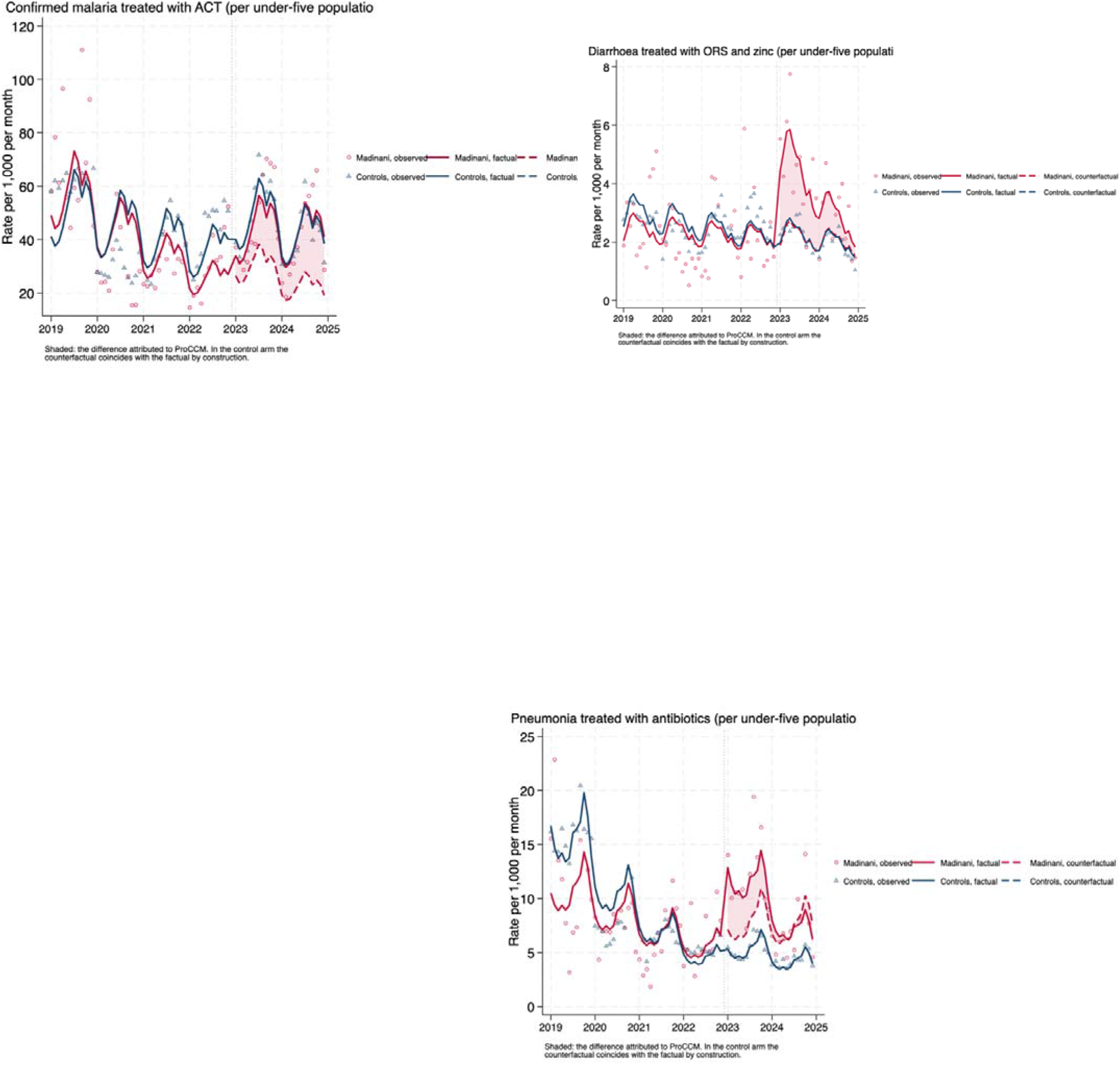
Monthly trend of child healthcare service utilization – facility level (Intervention versus control), from January 2019 to December 2024, with the ProCCM intervention implemented in December 2022 (vertical dashed line), created by authors.

**Supplementary Figure 9.**
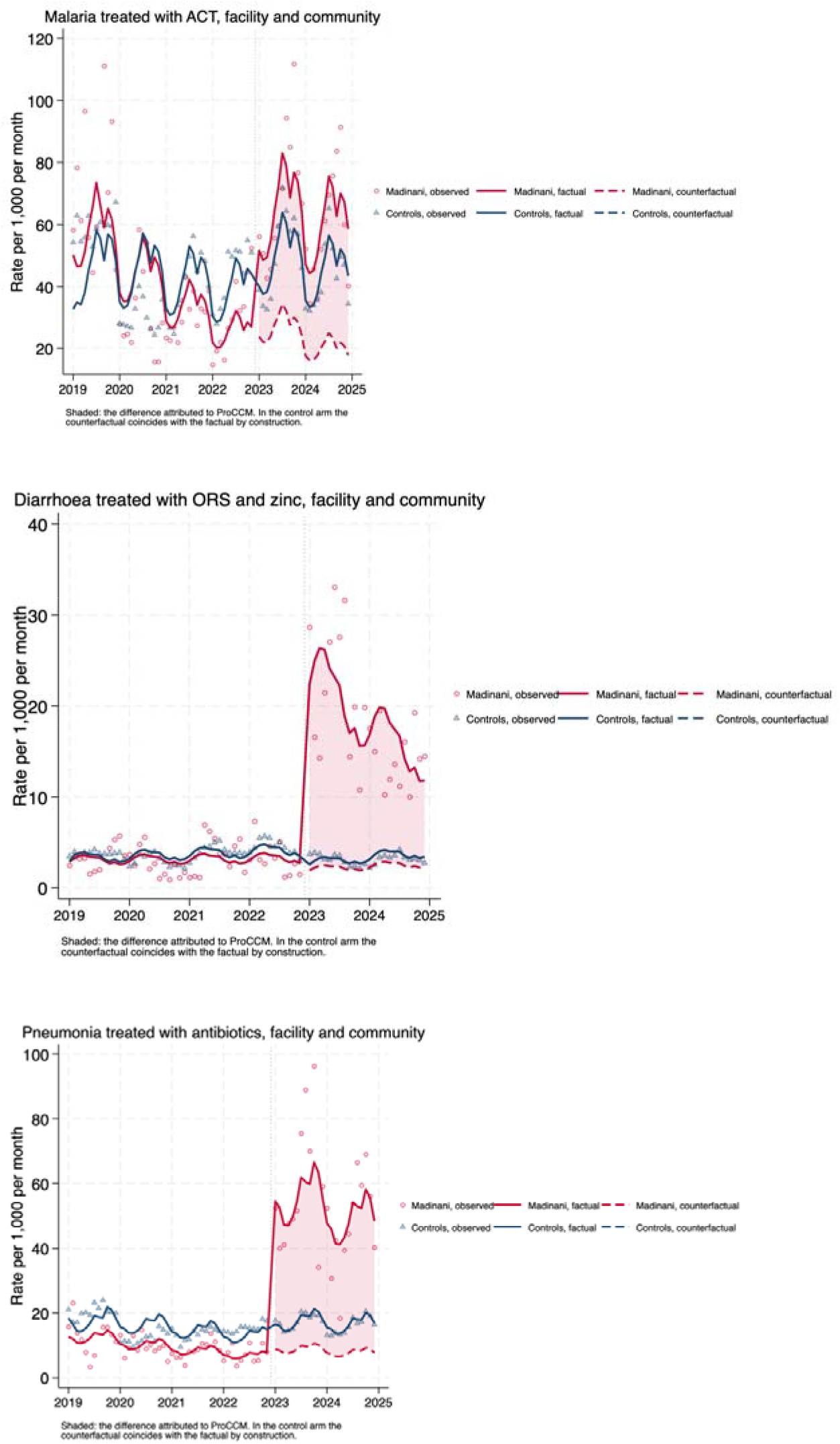
Monthly trend of child healthcare service utilization outcomes – facility and community level (Intervention versus control), from January 2019 to December 2024, with the ProCCM intervention implemented in December 2022 (vertical dashed line), created by authors.

**Supplementary Figure 10.**
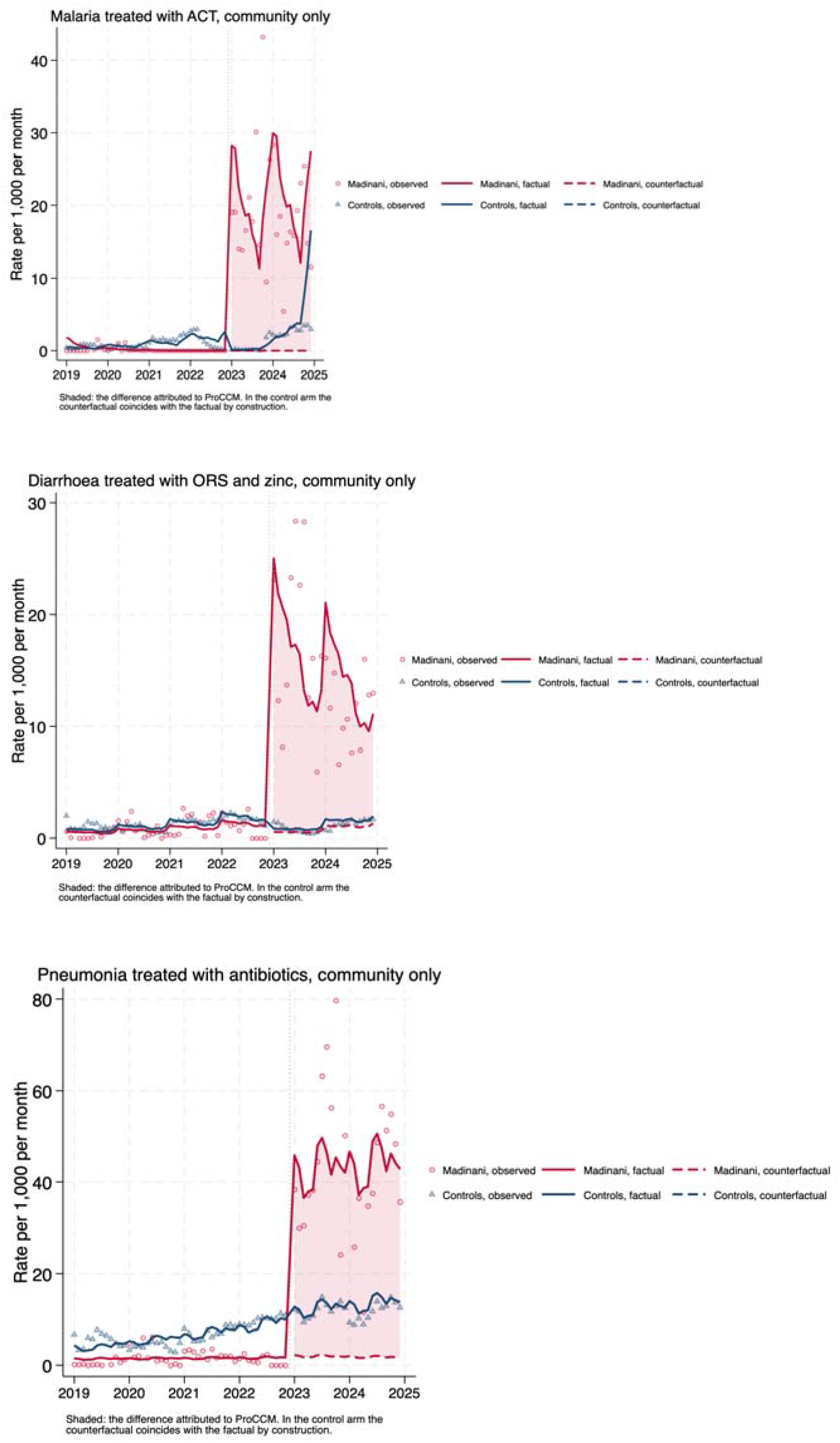
Monthly trend of child healthcare service utilization outcomes – community level (Intervention versus control), from January 2019 to December 2024, with the ProCCM intervention implemented in December 2022 (vertical dashed line), created by authors.

**Supplementary Figure 11.**
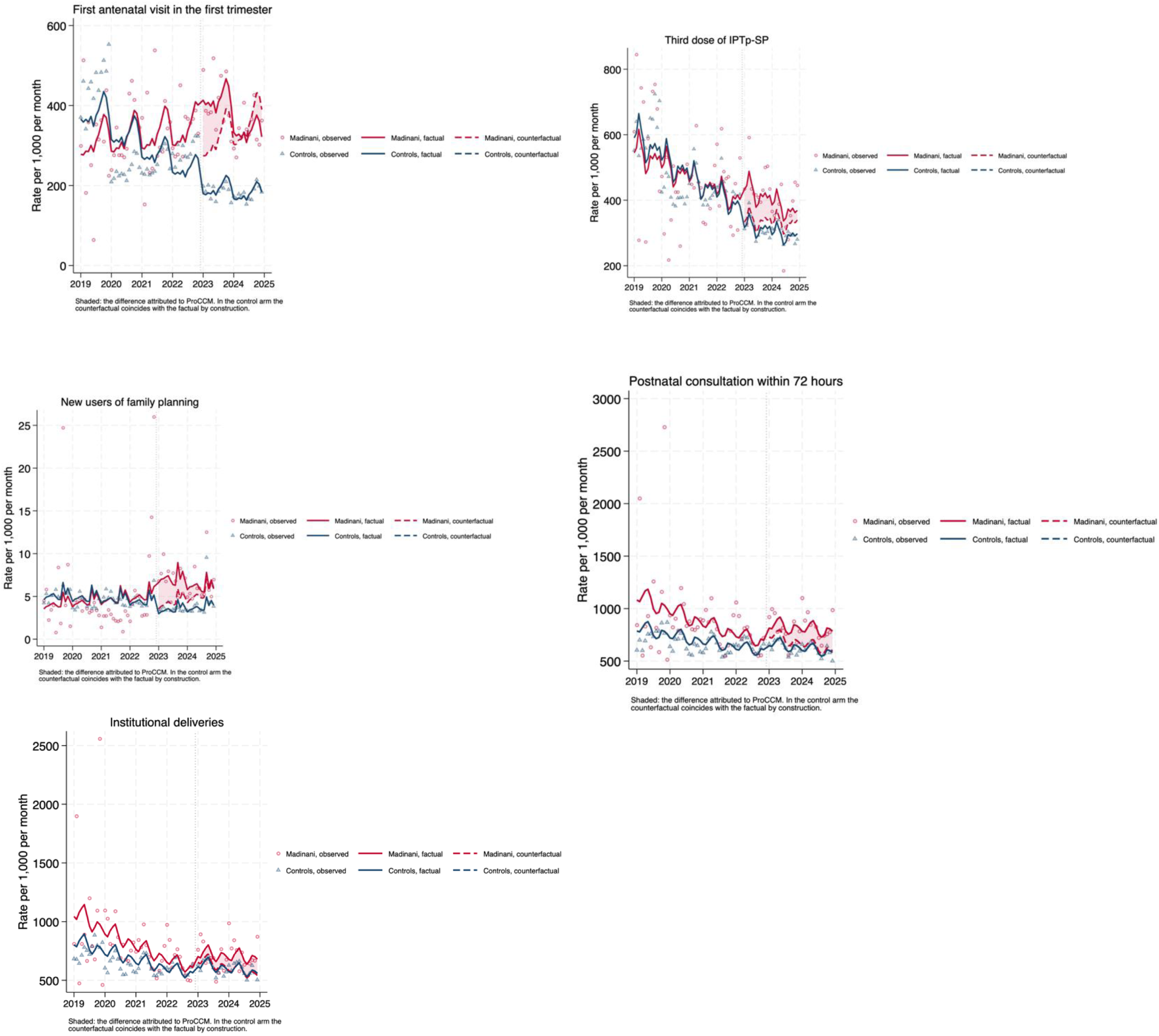
Monthly trend of maternal healthcare service utilization outcomes (Intervention versus control), from January 2019 to December 2024, with the ProCCM intervention implemented in December 2022 (vertical dashed line), created by authors.

**Supplementary Figure 12.**
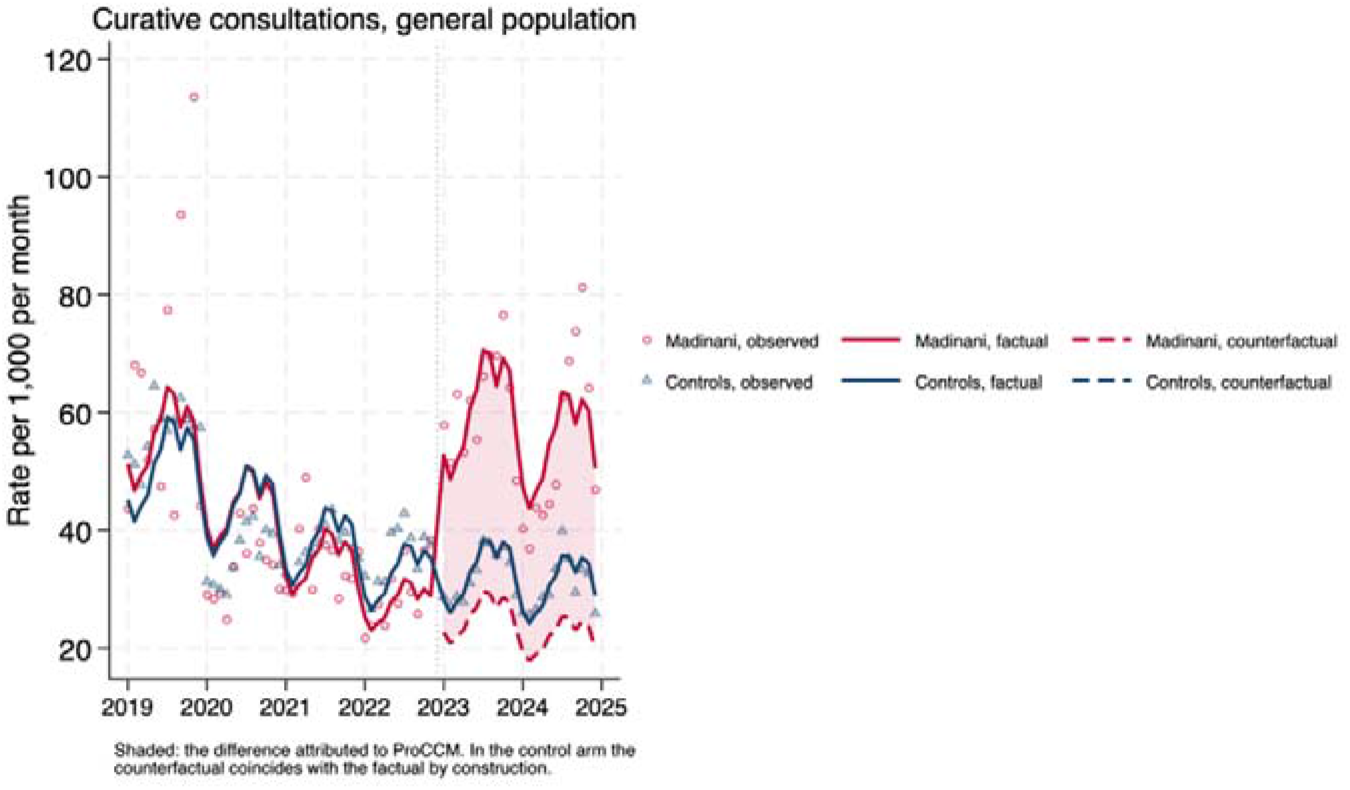
Monthly general curative consultations trend (Intervention versus control), from January 2019 to December 2024, with the ProCCM intervention implemented in December 2022 (vertical dashed line), created by authors.

**Supplementary Table S13.** Sensitivity analysis: level change (ratio of rate ratios) with comparison districts matched on the primary outcome, as in the main analysis, and re-selected on each outcome’s own pre-intervention series. Estimates are from a single dataset, so the first column differs slightly from Table 2.

|  | Estimated when the controls are fixed (matching based on the primary outcome of under-five consultations) |  |  |  | Estimated when the controls are re-selected on each outcome's own series |  |  |  |  |
|---|---|---|---|---|---|---|---|---|---|
|  | RRR | 95% CI |  | p | RRR | 95% CI |  | p | n_overlap |
| Under-five consultations, all causes | 2.00 | 1.57 | 2.54 | 0.00 | 2.00 | 1.57 | 2.54 | 0.00 | 20 |
| Curative consultations, general population | 2.20 | 1.79 | 2.71 | 0.00 | 2.08 | 1.69 | 2.56 | 0.00 | 13 |
| Confirmed malaria treated with ACT (per under-five consultation) | 0.63 | 0.50 | 0.80 | 0.00 | 0.62 | 0.52 | 0.74 | 0.00 | 6 |
| Diarrhoea treated with ORS and zinc (per under-five consultation) | 0.94 | 0.62 | 1.42 | 0.77 | 0.96 | 0.69 | 1.34 | 0.82 | 6 |
| Pneumonia treated with antibiotics (per under-five consultation) | 0.75 | 0.54 | 1.04 | 0.09 | 0.95 | 0.75 | 1.21 | 0.70 | 2 |
| First antenatal visit in the first trimester | 1.40 | 1.00 | 1.95 | 0.05 | 2.01 | 1.65 | 2.46 | 0.00 | 5 |
| Third dose of IPTp-SP | 1.36 | 1.07 | 1.73 | 0.01 | 1.32 | 1.05 | 1.67 | 0.02 | 8 |
| Institutional deliveries | 1.01 | 0.77 | 1.33 | 0.93 | 1.06 | 0.90 | 1.24 | 0.51 | 9 |
| Postnatal consultation within 72 hours | 1.03 | 0.79 | 1.36 | 0.82 | 1.09 | 0.93 | 1.28 | 0.30 | 8 |
| New users of family planning | 1.94 | 1.30 | 2.89 | 0.00 | 1.82 | 1.31 | 2.52 | 0.00 | 6 |
| Confirmed malaria treated with ACT (per under-five population) | 1.25 | 0.88 | 1.77 | 0.22 | 1.26 | 0.91 | 1.75 | 0.17 | 12 |
| Diarrhoea treated with ORS and zinc (per under-five population) | 1.92 | 1.24 | 2.96 | 0.00 | 2.09 | 1.45 | 3.00 | 0.00 | 6 |
| Pneumonia treated with antibiotics (per under-five population) | 1.54 | 1.05 | 2.27 | 0.03 | 2.07 | 1.51 | 2.83 | 0.00 | 10 |
| Malaria treated with ACT, facility and community | 1.87 | 1.34 | 2.63 | 0.00 | 1.90 | 1.39 | 2.60 | 0.00 | 12 |
| Diarrhoea treated with ORS and zinc, facility and community | 8.38 | 5.39 | 13.02 | 0.00 | 9.30 | 6.36 | 13.60 | 0.00 | 7 |
| Pneumonia treated with antibiotics, facility and community | 4.97 | 3.39 | 7.27 | 0.00 | 5.55 | 4.05 | 7.59 | 0.00 | 11 |
| Malaria treated with ACT, community only |  |  |  |  |  |  |  |  |  |
| Diarrhoea treated with ORS and zinc, community only | 55.53 | 16.65 | 185.23 | 0.00 | 57.17 | 21.27 | 153.64 | 0.00 | 4 |
| Pneumonia treated with antibiotics, community only | 23.12 | 8.98 | 59.52 | 0.00 | 28.17 | 14.84 | 53.50 | 0.00 | 4 |

n_overlap: number of comparison districts shared with those selected on the primary outcome. p shown as 0.00 indicates p < 0.005. Community-level malaria is not shown because Madinani's pre-intervention count was close to zero.

**Supplementary Table S14.**
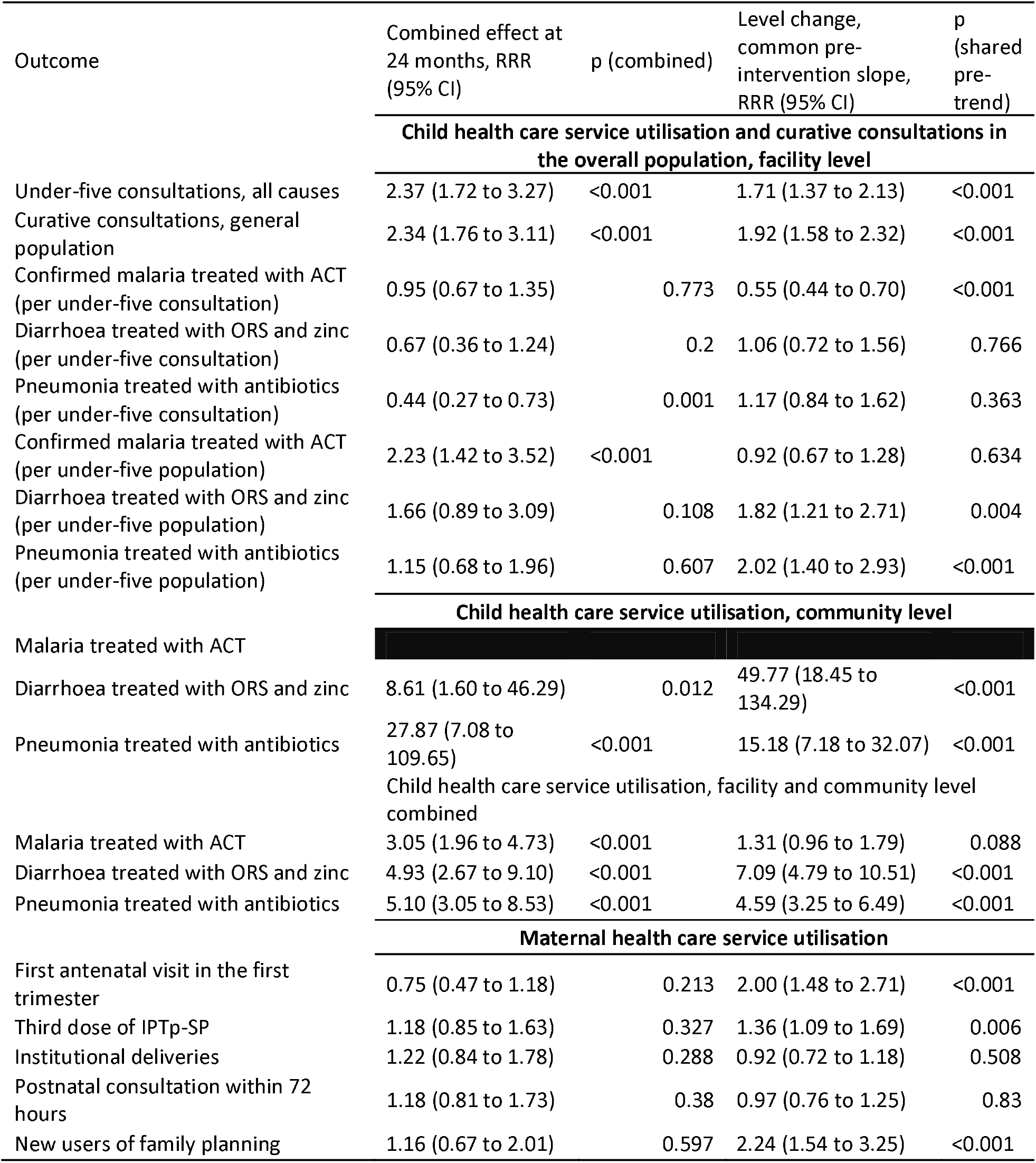
Combined effect 24 months after launch (primary specification) and level change under a common pre-intervention slope, as ratios of rate ratios. Community-level malaria is not shown (see Table 2).

**Supplementary Figure 15.**
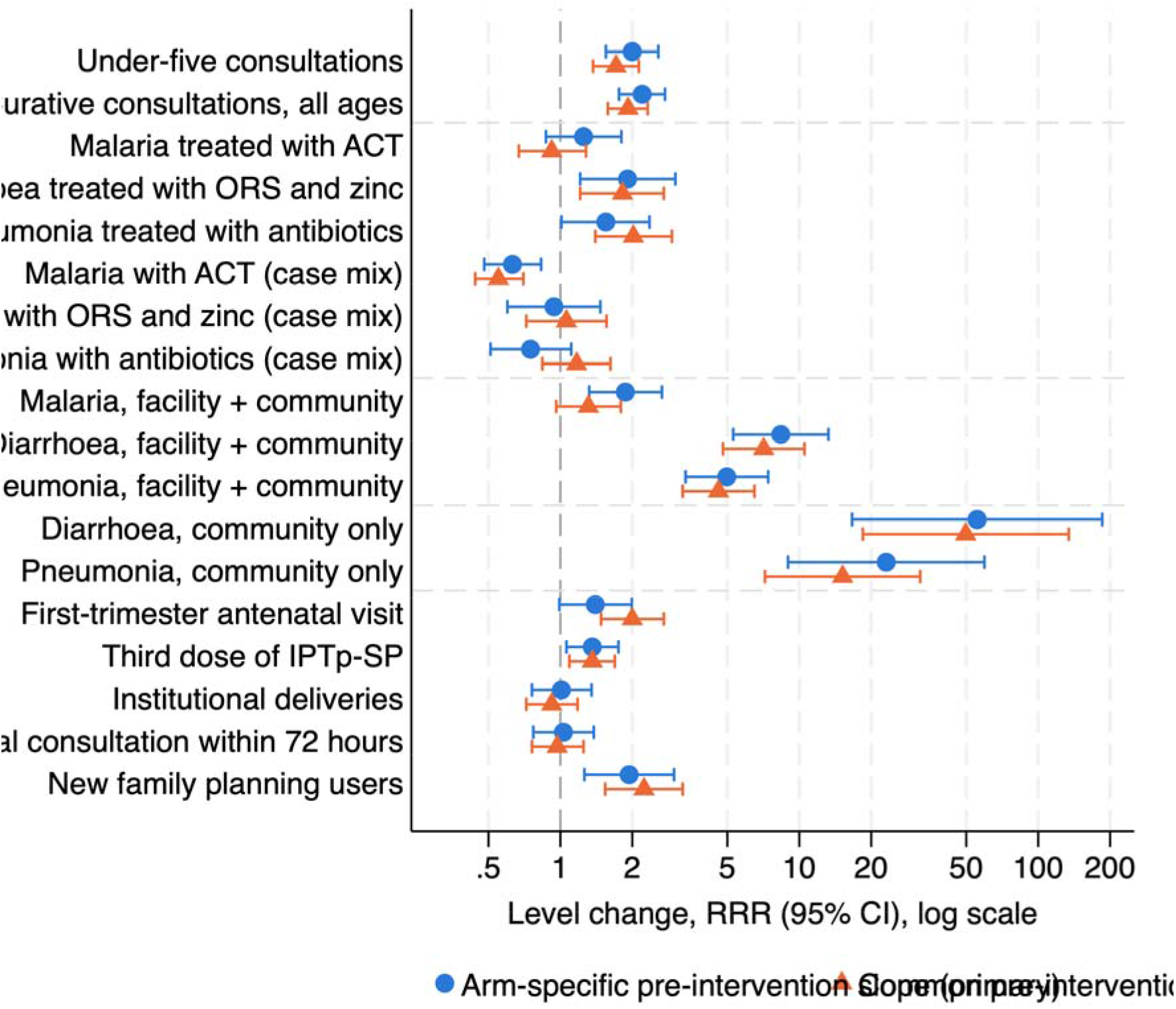
Level change for each outcome under the primary specification (arm-specific pre-intervention slope) and under a common pre-intervention slope (ratio of rate ratios, 95% CI).

**Supplementary Figure 16.**
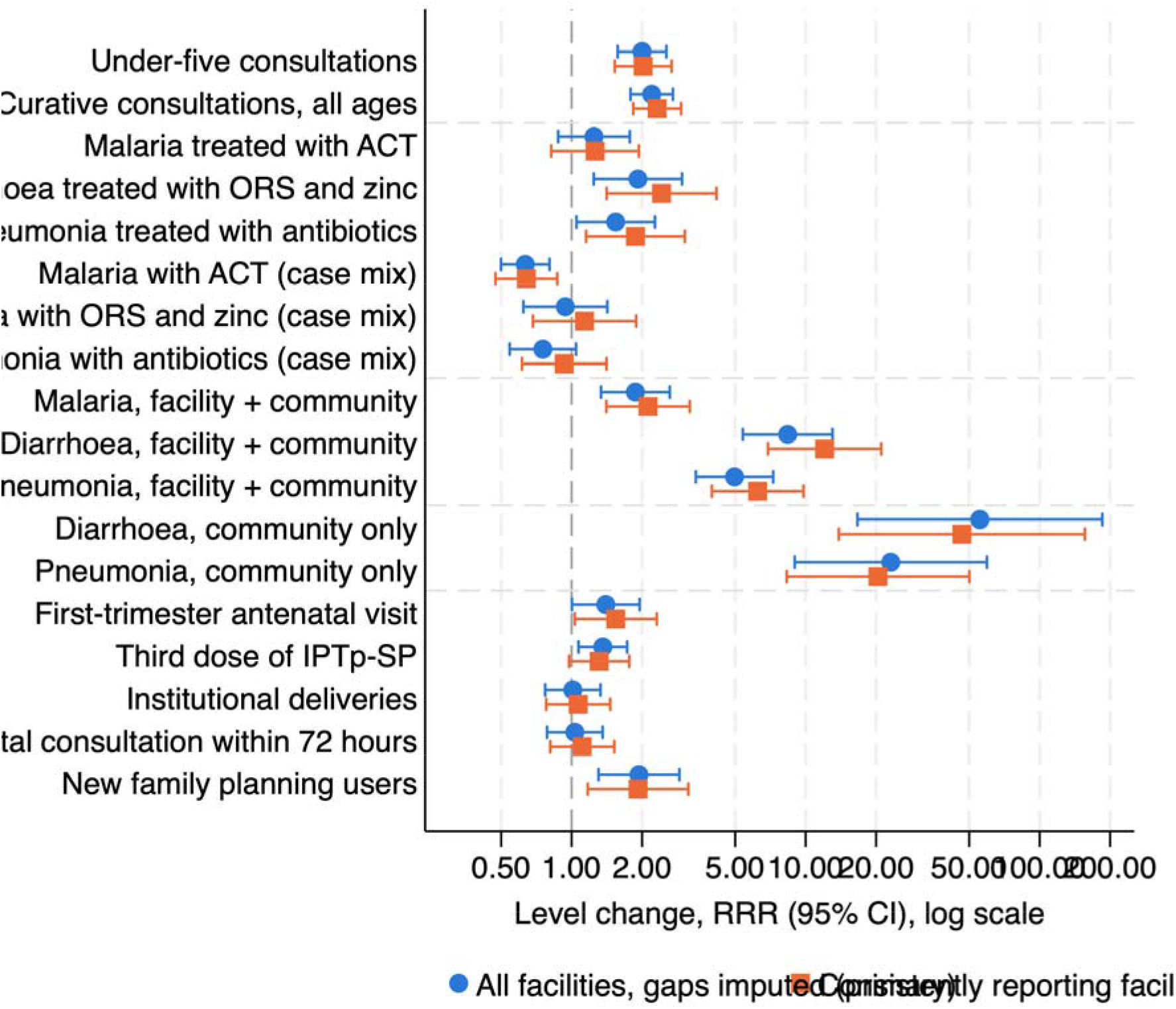
Level change variations according to completeness of data (Imputed versus nearly complete facilities only), created by authors.

**Supplementary Table S17.** Sensitivity analysis with district-specific random slopes on calendar time: level change under the primary model and under two random-slope models (Madinani’s slope estimated separately, or partially pooled with the comparison districts’ slopes), change in slope at launch, and Madinani’s pre-intervention slope. All models in this table were fitted to a single dataset, so the primary-model column differs slightly from Table 2.

| Outcome | Primary model: level change, RRR (95% CI) | Random slopes, Madinani slope separate: level change | Random slopes, Madinani slope partially pooled: level change | Change in slope at launch, RRR per month (95% CI), random slopes | Madinani pre-intervention slope deviation (SD of district slopes) | Rank of Madinani's partially pooled pre-intervention slope (1 = steepest decline, of 21) |
|---|---|---|---|---|---|---|
| Under-five consultations (facility) | 2.00 (1.57 to 2.54) | 2.01 (1.61 to 2.52) | 1.94 (1.56 to 2.42) | 1.008 (0.994 to 1.021) | -1.67 | 3 |
| General-population consultations | 2.20 (1.79 to 2.71) | 2.21 (1.82 to 2.68) | 2.15 (1.78 to 2.59) | 1.003 (0.991 to 1.015) | -1.58 | 4 |
| Malaria treated with ACT (facility) | 1.25 (0.88 to 1.77) | 1.27 (0.92 to 1.75) | 1.20 (0.87 to 1.66) | 1.026 (1.006 to 1.046) | -1.95 | 3 |
| Diarrhoea, ORS and zinc (facility) | 1.92 (1.24 to 2.96) | 2.03 (1.40 to 2.96) | 2.01 (1.39 to 2.91) | 0.997 (0.974 to 1.020) | -0.38 | 10 |
| Pneumonia, antibiotics (facility) | 1.54 (1.05 to 2.27) | 1.61 (1.15 to 2.25) | 1.65 (1.19 to 2.30) | 0.989 (0.969 to 1.010) | 1.14 | 20 |
| Malaria treated with ACT (facility + community) | 1.87 (1.34 to 2.63) | 1.93 (1.41 to 2.64) | 1.83 (1.34 to 2.49) | 1.022 (1.003 to 1.042) | -2.42 | 2 |
| Diarrhoea, ORS and zinc (facility + community) | 8.38 (5.39 to 13.02) | 8.87 (6.08 to 12.94) | 8.66 (5.97 to 12.57) | 0.983 (0.961 to 1.005) | -0.82 | 5 |
| Pneumonia, antibiotics (facility + community) | 4.97 (3.39 to 7.27) | 5.21 (3.73 to 7.29) | 5.13 (3.69 to 7.14) | 1.004 (0.984 to 1.025) | -0.63 | 8 |
| First-trimester antenatal care | 1.40 (1.00 to 1.95) | 1.42 (1.05 to 1.91) | 1.50 (1.12 to 2.01) | 0.976 (0.959 to 0.994) | 2.21 | 21 |
| Third-dose IPTp-SP | 1.36 (1.07 to 1.73) | 1.37 (1.13 to 1.67) | 1.37 (1.13 to 1.66) | 0.996 (0.984 to 1.007) | -0.08 | 10 |
| New family planning users | 1.94 (1.30 to 2.89) | 1.94 (1.32 to 2.85) | 2.03 (1.41 to 2.94) | 0.979 (0.956 to 1.002) | 1.14 | 17 |

**Supplementary Table S18.** Pre-intervention trend test: ratio of Madinani’s monthly pre-intervention trend to that of the 20 matched comparison districts (arm-by-time interaction fitted to pre-intervention data only). p < 0.05 indicates non-parallel pre-intervention trends.

| Outcomes | Pre-intervention trend ratio | 95% CI Lower | 95% CI Higher | p-value |
|---|---|---|---|---|
| Under-five consultations, all causes | 0.994 | 0.989 | 0.998 | 0.010 |
| Curative consultations, general population | 0.994 | 0.990 | 0.999 | 0.012 |
| Confirmed malaria treated with ACT (per under-five consultation) | 0.994 | 0.989 | 1.000 | 0.036 |
| Diarrhoea treated with ORS and zinc (per under-five consultation) | 1.004 | 0.995 | 1.014 | 0.346 |
| Pneumonia treated with antibiotics (per under-five consultation) | 1.019 | 1.012 | 1.026 | 0.000 |
| First antenatal visit in the first trimester | 1.015 | 1.008 | 1.023 | 0.000 |
| Third dose of IPTp-SP | 1.000 | 0.995 | 1.004 | 0.898 |
| Institutional deliveries | 0.996 | 0.989 | 1.002 | 0.205 |
| Postnatal consultation within 72 hours | 0.997 | 0.991 | 1.004 | 0.453 |
| New users of family planning | 1.005 | 0.997 | 1.014 | 0.206 |
| Confirmed malaria treated with ACT (per under-five population) | 0.988 | 0.981 | 0.995 | 0.001 |
| Diarrhoea treated with ORS and zinc (per under-five population) | 0.997 | 0.988 | 1.006 | 0.508 |
| Pneumonia treated with antibiotics (per under-five population) | 1.011 | 1.003 | 1.019 | 0.006 |
| Malaria treated with ACT, facility and community | 0.985 | 0.978 | 0.992 | 0.000 |
| Diarrhoea treated with ORS and zinc, facility and community | 0.992 | 0.983 | 1.001 | 0.065 |
| Pneumonia treated with antibiotics, facility and community | 0.996 | 0.988 | 1.004 | 0.280 |
| Malaria treated with ACT, community only | 0.832 | 0.781 | 0.887 | 0.000 |
| Diarrhoea treated with ORS and zinc, community only | 0.990 | 0.964 | 1.016 | 0.444 |
| Pneumonia treated with antibiotics, community only | 0.981 | 0.954 | 1.008 | 0.165 |

